# The Effects of a Multidomain Lifestyle Intervention on Brain Function and Its Relation with Immunometabolic Markers and Intestinal Health in Older Adults at Risk of Cognitive Decline: The HELI Randomised Controlled Trial

**DOI:** 10.64898/2026.07.28.26358647

**Authors:** Mark R. van Loenen, Nienke R.K. Zwart, Lianne B. Remie, Mara P.H. van Trijp, Michelle G. Jansen, José P. Marques, Jurgen A.H.R. Claassen, Mechteld M. Grootte Bromhaar, Ondine van de Rest, Inge M.W. Verberk, Yannick Vermeiren, Guido J. Hooiveld, Wilma Steegenga, Nynke Smidt, Sietske A.M. Sikkes, Kay Deckers, Marissa D. Zwan, Sebastian Köhler, Joukje M. Oosterman, Esther Aarts

## Abstract

**Introduction:** Multidomain lifestyle interventions may slow down cognitive decline and dementia, but the brain mechanisms underlying these effects remain unclear. We investigated brain, inflammation, cardiometabolic, and gut health changes associated with a six-month multidomain lifestyle intervention in Dutch older adults at risk for cognitive decline and dementia.

**Method:** Participants aged 60–75 years with ≥2 lifestyle-modifiable cardiovascular risk factors (e.g., BMI ≥25, physical inactivity) were recruited and randomised in a 1:1 ratio to a high-intensity intervention or active control group using stratified (2,4) block-randomisation. The multidomain lifestyle intervention included five lifestyle domains (diet, physical activity, stress management and mindfulness, cognitive training, and sleep). Primary outcomes were changes in cerebral perfusion levels, brain activity during working memory, working memory performance, inflammation profile, and microbiota diversity. Secondary outcomes included lifestyle factors and markers of neuroimaging, brain health, cardiovascular and gut health, and cognitive functioning. The effects of the lifestyle intervention were investigated using linear mixed-effect models.

**Results:** A total of 102 participants were randomised into the intervention (n=53) or active control group (n=49), of which 86 participants completed the six-month multidomain lifestyle intervention. Baseline characteristics were similar between the groups. The intervention resulted in improved diet and sleep scores in the intervention group compared to the active control group (Beta 1.67, 95% CI 0.61;2.73, PFDR=0.01, and Beta −1.80, 95% CI −2.79;-0.81, PFDR=0.005, respectively). Within the intervention group, favourable reductions in lifestyle-modifiable cardiovascular risk factors, such as BMI and blood pressure, were observed, but these were not significantly different from changes in the active control group. The intervention did not result in significant changes over time between the intervention and control group in other primary and secondary outcomes.

**Conclusion:** The HELI multidomain lifestyle was able to reduce lifestyle-modifiable cardiovascular risk factors associated with dementia, although not significant compared to the active control group. We did not observe significant intervention effects on brain, cardiometabolic, inflammatory, and gut health outcomes. These results emphasize the complexity of exploring the role of lifestyle on neurobiological brain mechanisms associated with maintaining optimal cognitive functioning in aging and their underlying peripheral (to central) mechanisms.

**Clinical Trial Registration Information:** The HELI study was approved by an accredited Dutch national research ethics committee, the Medical Research Ethics Committee (MREC) Oost-Nederland (file number NL78263.091.21). This study is registered with ClinicalTrials.gov (Trial ID NCT05777863).

## Introduction

The number of people affected by cognitive decline and dementia has been steadily increasing globally over the past decades due to population greying [1, 2]. The Lancet commission on dementia reported fourteen modifiable risk factors potentially affecting disease course for about 45% of global dementia cases [2, 3], highlighting the opportunities for dementia risk reduction. These risk factors include lifestyle-related factors such as obesity, physical inactivity, hypertension, hypercholesterolemia, and type-II diabetes [4–6]. Interventions targeting multiple different domains of lifestyle simultaneously, including diet, physical activity, stress management, and other health behaviours, hold great potential to address the diverse established risk factors contributing to dementia.

In recent years, multiple studies have investigated the efficacy of multidomain lifestyle interventions in reducing risk profiles of dementia, such as the FINGER and US POINTER studies [7–13]. A meta-analysis further reported that multiple multidomain lifestyle interventions in aging populations improved cognition and reduced dementia risk, especially in subgroups with increased risk of cognitive decline and dementia, e.g., elderly individuals with cardiovascular risk factors [14]. However, several recent multidomain lifestyle intervention trials have reported no effects on cognition [15, 16]. In addition, among studies demonstrating cognitive benefits, effect sizes were small. Collectively, these studies have reported varying results, have large differences in follow-up time, ranging from three months to up to six years, and none have demonstrated a reduction in dementia incidence. One way to better understand these data and the small effect sizes reported, is to explore changes in the possible underlying brain and peripheral mechanisms that potentially drive the reported cognitive benefits of multidomain lifestyle interventions.

We, therefore, set-up the HELI (“Hersenfuncties na Leefstijl Interventie”) study. The overall aim of the HELI study is to investigate the brain mechanisms of a six-month multidomain lifestyle intervention and their underlying peripheral (to central) mechanisms in a cohort of older adults who had several lifestyle and cardiovascular risk factors associated with dementia incidence [17]. Our primary objectives were to evaluate intervention effects on (1) cerebral perfusion levels, (2) brain activity during working memory, (3) working memory performance, (4) inflammatory profile in blood, and (5) microbiota diversity. Secondary objectives were to (1) investigate intervention effects on additional neuroimaging and biomarkers of brain health, lifestyle domain adherence, cardiovascular and gut health, and cognitive functioning, and (2) to explore whether changes in (primary) brain outcomes were correlated with changes in (primary) peripheral outcomes. With this mechanism-oriented study, we aim to clarify why previous multidomain lifestyle intervention studies have shown small and mixed effects on cognitive benefits and to improve our understanding of the subtle effects and individual differences underlying these outcomes.

## Methods

### Study design

The HELI study was a single-blinded, two-armed parallel randomised controlled trial with a high-intensity group (intervention group) and a low-intensity group (active control group). Details of the design of the study have been previously described [17]. The study was approved by a Medical Research Ethics Committee (MREC Oost-Nederland NL78263.091.21). All participants provided written informed consent.

### Eligibility criteria

The HELI study included Dutch participants aged 60–75 years, with presence of lifestyle-modifiable cardiovascular risk factors, included in the Cardiovascular Risk Factors, Aging and Incidence of Dementia (CAIDE) risk index [18]. Participants were considered eligible if they self-reported two or more of the following: overweight (body mass index (BMI) ≥25 kg/m^2^), physical inactivity (according to 2020 WHO guidelines [19]), hypertension (systolic blood pressure ≥140 mmHg and diastolic blood pressure ≥90 mmHg), hypercholesterolemia (total cholesterol >5 mmol/L and/or LDL-cholesterol >3 mmol/L), diabetes type-II, or mild cardiovascular disease (e.g., intermittent claudication, varicose veins). Hypertension contributed one risk point with an additional point assigned if participants were not using antihypertensive medication. Participants were excluded if they provided self-reported indications of the following: cognitive impairment or dementia, not fluent in spoken and written Dutch, contraindications for magnetic resonance imaging (MRI), or clinical conditions that could interfere with intervention participation or study outcomes. The modified Telephone Interview for Cognitive Status (TICS-M1) was additionally performed to exclude participants with undiagnosed cognitive impairment or dementia as determined by a score <23 [20]. The full inclusion and exclusion criteria and full pre-screening procedure have been described in detail elsewhere [17]. All participants were fully informed about the purpose, procedures, and potential risks of the study. Participation was voluntary and participants could withdraw from the study at any time.

### Randomisation and masking

We utilized a stratified (2,4) block-randomisation based on sex (female/male), inclusion risk factor profile (medium risk: 2–3 points/high risk ≥4 points), and a combined factor of education level and age (≥70 years or low education/60-69 years and high education). Age and education were combined in one stratification factor as they are both important covariates of cognitive functioning and cognitive decline [21]. Education level was classified as low, medium, or high according to the 2011 International Standard Classification of Education (ISCED) [16]. An independent researcher allocated participants to the two intervention arms: a low-intensity general health advice group (active control) and a high-intensity supervised group coaching intervention group. This study was single-blind, as participants were aware of the intervention arm assignment. Due to ethical considerations, intervention content had to be disclosed before formal inclusion, making blinding of the intervention impossible. Outcome measure visits and data acquisition were conducted by researchers blinded to intervention allocation and all data preprocessing and analyses were performed using dummy participant IDs. A more extensive description of study randomisation and blinding has been reported previously [13].

### Intervention

An extensive description of the intervention group design, lifestyle domain-specific contents and rationale has been previously described [17]. In short, the six-month HELI multidomain lifestyle intervention was based on the FINGER-NL [22] intervention design, which is part of the World-Wide FINGERS initiative [23]. The intervention targeted five different lifestyle domains: (1) diet, (2) physical activity, (3) stress management and mindfulness, (4) cognitive training, and (5) sleep. The active control group received general lifestyle-related health information on each of the lifestyle domains, through e-mail once every two weeks. The intervention group followed a structured and supervised intervention program with weekly group meetings, exercises, and course-program materials. The weekly intervention group meetings were predominantly held online. Participants received lifestyle-related information, advice, and exercises about each of the five incorporated lifestyle domains.

### Data collection

#### Lifestyle questionnaires

At baseline and after the six-month intervention, participants filled in questionnaires to assess adherence to various lifestyle factors. The lifestyle questionnaires used are previously described in detail [24]. In short, lifestyle-related questionnaires included the Eetscore food frequency questionnaire to assess adherence to the 2015 Dutch dietary guidelines (Dutch Healthy Diet; DHD) index [25] and to the Dutch MIND (MIND-NL [26, 27]) criteria; the Short Questionnaire to Assess Health-enhancing physical activity (SQUASH [28]) as a measure for physical activity; the LASA Sedentary Behaviour Questionnaire (SBQ [29]) to quantify weekly sedentary behaviour; the Perceived Stress Scale (PSS [30]) to measure situational stress perception; the Hospital Anxiety and Depression Scale (HADS [31]) to assess depressive and anxiety symptoms; the Five Facet Mindfulness Questionnaire (FFMQ [32]) to assess multiple subscales of mindfulness (e.g., non-judgmental thoughts, acting with awareness); and the Pittsburgh Sleep Quality Index (PSQI [33]) to assess multiple components of sleep quality. For the HADS, FFMQ and PSQI, we calculated a total score by summing the individual scores of their respective subscales or components.

#### Anthropometric measurements

For anthropometric measurements, we used BMI, waist-hip ratio, and blood pressure. Height and weight were measured to calculate BMI (kg/m^2^). Waist and hip circumference were measured using a tape measure at baseline and six-month follow-up. Systolic blood pressure (SBP) and diastolic blood pressure (DBP) were measured twice with an Omron X3 Comfort digital blood pressure monitor, with a third measurement obtained if the first two showed substantial variation. All available SBP and DBP measurements were subsequently averaged. The averaged blood pressure values were then used to calculate mean arterial pressure (MAP) as DBP + ⅓ × (SBP − DBP) [34].

#### Neuroimaging

For neuroimaging measurements, we obtained MRI scans with a Siemens (Erlangen, Germany) 3T MAGNETOM Skyra MR scanner at the Donders Centre for Cognitive Neuroimaging, Radboud University, in Nijmegen, the Netherlands. First baseline MRI scans started in May 2022, and final follow-up scans were concluded in May 2024. The neuroimaging scanning protocol included a (1) structural T1-weighted Magnetization Prepared 2 Rapid Acquisition Gradient Echoes (MP2RAGE) sequence (TR/TE=6000/2.34 ms, TI1/TI2=700/2400 ms, voxel size=1.0 mm isotropic, flip angle=6/6°); (2) N-back task-based fMRI (TR/TE=1500/33.40 ms, voxel size=20 mm isotropic, flip angle=75°) EPI sequence; (3) pseudocontinuous arterial spin labelling (pCASL) (TR/TE=4000/16.28 ms, voxel size=1.8×1.8×3.5 mm, flip angle exc/ref=90/120°); and a (4) Point Resolved Spectroscopy (PRESS) Magnetic Resonance Spectroscopy (MRS) sequence (TR/TE=2000/35 ms, voxel size=20 mm isotropic, flip angle=90×180×180°). A detailed overview of all MRI sequence parameters has been provided previously [17].

For fMRI measurements, we used an N-back task to investigate working memory-related neural recruitment and task performance. Participants viewed digits (1-9) and responded when the stimulus matched one presented n trials earlier (n=0, 1, or 2). The 0-back served as a control condition, only requiring attention (respond to “1”). The block-design task lasted 15 minutes and participants practiced beforehand to ensure understanding. Due to a necessary MRI scanner update in 2023 during data acquisition, the pCASL sequence was changed. Although the sequence parameters remained identical, the sequence version (pre-vs post-update) was included as a covariate in the statistical models including cerebral blood flow (CBF) measures to account for potential sequence-specific effects. An extensive overview of all sequence parameters, including parameters for calibration images and B1 maps, has been provided previously [17]. See **Supplementary File 1** for the method on imaging preprocessing and analysis.

#### Cognitive assessment

We used a neuropsychological test battery to assess cognitive domains predominantly affected by cognitive decline. This included the Trail Making Test (TMT) [35] part A and B (TMT-A/B) and Verbal Fluency Test (VFT) [36] for executive functioning; the Digit Symbol Substitution Test (DSST) [37] for processing speed; the forward and backward-summed score of the Digit Span Test (DST) [37] and N-back task performance (during fMRI) for working memory; and the delayed-recall Rey Auditory Verbal Learning Test (RAVLT) [38] for episodic memory. To account for TMT-A completion time, we calculated a TMT-B/TMT-A ratio score. We used the VFT ‘Animals’ at baseline and follow-up. For the DST, we calculated a sum-score of the forward and backward DST scores. N-back task performance was measured with the d-prime (d’), calculated as d’ = z(H) - z(F) where H is the hit rate, and F is the false alarm rate.

#### Blood markers

Blood samples were collected at baseline and six-month follow-up to analyse inflammatory, cardiometabolic, intestinal integrity, and neurodegeneration (brain health) markers. Blood samples were collected in the morning and participants were instructed to remain fasted from 20.00 hrs the day before the visit. Blood was drawn via finger prick and intravenously from the arm. Blood collected intravenously was used to analyse cardiometabolic markers (glucose, insulin, cholesterol, total-high-density lipoprotein (HDL) cholesterol, total-low-density lipoprotein (LDL) cholesterol, and triglycerides all in mmol/L), inflammation markers (IL-6, TNF-α, IFN-γ, IL-8, and IL-10 all in pg/ml), intestinal integrity markers (lipopolysaccharide (LPS) in EU/ml, lipopolysaccharide binding protein (LBP) in µg/ml, and zonulin in ng/ml), and blood biomarkers of neurodegeneration (neurofilament light chain (NfL), amyloid-β 42 (Aβ42), amyloid-β 40 (Aβ40), and glial fibrillary acidic protein (GFAP) all in pg/ml). Blood tubes were inverted ten times and afterwards centrifuged (3000 × g) for 8 min at 20°C. Plasma and serum were collected and stored at −80°C until further analysis.

The methods for cardiometabolic marker analyses are previously described [24]. Inflammation markers were quantified using a mesoscale discovery V-PLEX Pro inflammatory Panel 1 (human) kit (K151A9H-1, Mesoscale Discovery, Gaithersburg, MD) at Wageningen University and Research according to the manufacturer’s instructions. ELISAs were used to quantify zonulin and LPS-binding protein (LBP) in 25 µL of serum each. Zonulin was measured using the K5601 assay kit (Immundiagnostik AG, Bensheim, Germany) and LBP was measured using the HK315-02 assay kit (Hycult Biotech, Uden, Netherlands), at Wageningen University and Research following manufacturers’ instructions. LPS was measured in serum with a chromogenic assay from Nodia (associates Cap. Cod Inc.) at Wageningen University and Research. The assay was performed in a Pyros Kinetix Flex tube reader (pKFlex) (Nodia, Cap. Cod Inc.). Pyrochrome lysate was reconstituted with 3.4 ml Pyrochrome reconstitution buffer (C1500-5) and for the calibration curve Limulus Amebocyte lysate control standard with an endotoxin concentration of 0.5 µg/vial (CSEE0005-1) was used. The samples were diluted with LPS free MQ and incubated for 15 min in a water bath at 70 °C. After 1 h at 4 °C the samples were placed for 15 min at room temperature and directly measured on the pKflex. 200 µl treated sample or standard was added in a pKFlex glass tube, 50 µl Pyrochrome was added, and the mixture was shortly stirred and placed in the pKFlex. Using the Pyrosexpress 21 CFR Part 11 compliant software the endotoxin concentration was calculated. All samples were analysed in duplicate. For the analysis of the biomarkers reflective of neurodegeneration-related processes, samples were shipped to Amsterdam UMC, Laboratory Medicine, Neurochemistry laboratory on dry ice. Analyses were performed using the Simoa neurology 4-plex E assay kit (Quanterix, Billerica, United States of America) on the Simoa HD-X analyser. Analyses were performed in monoplo, with on-board 4-times sample dilution according to manufacturers’ instructions [39]. Prior to analysis, samples were shortly thawed at room temperature in front of a cold-air fan, and subsequently vortexed and centrifuged at 10,000xg for 10 minutes.

C-reactive protein (CRP) levels and white blood cell (WBC) counts were determined from blood droplets collected via finger prick. For CRP concentration measurement, 20 µL of blood was collected in a capillary tube, directly transferred into a cuvette with buffer solution, and analysed using the QuikRead Go analyser (Orion Diagnostics, Ghodbunder, India) according to the manufacturer’s instructions. The measurement range was between 0.5–200 mg/L. For total white blood cell counts and differential counts of neutrophils, lymphocytes, monocytes, eosinophils, and basophils, 10 µL of blood was collected in a microcuvette, and analysed using the HemoCue® WBC DIFF (HemoCue AB, Ängelhom, Sweden) following manufacturer’s instructions. The measurement range was between 0.3-30.0×10^9^/L.

#### Microbiota composition

At baseline and six-month follow-up, faecal samples were collected. Faecal samples were processed under frozen conditions in two phases to minimize variability in gut health markers [40]. First, the faeces were crushed into smaller particles (∼0.5-1 cm²) using a dead-blow hammer (Performance Tool Dead-Blow Hammer, Wilmar Corporation) while kept frozen with dry ice and liquid nitrogen. The hammered samples were then stored at −80°C. Second, a portion of the crushed faeces was further processed using the IKA® Tube Mill 100 control (IKA-Werke GmbH & Co. KG, Staufen, Germany). Approximately 10 grams of the crushed sample was placed in a 40 mL disposable milling chamber (art. IKAA20001173, IKA) along with two pieces of dry ice. Faeces were milled under frozen conditions for approximately 20 sec at 25000 rpm, pulverizing it into a fine, homogeneous powder. The homogenized faeces were stored at −80°C until further analysis.

Bacterial DNA was extracted from approximately 250 mg faeces using the DNeasy® PowerSoil® Pro Kit (QIAGEN, Hilden, Germany), following the manufacturer’s instructions. Cell lysis and bead beating were performed using the TissueLyser adapter set 2 (QIAGEN, Hilden, Germany). DNA concentrations were quantified using a Nanodrop ND-1000 spectrophotometer (Thermo Fisher Scientific, Waltham, MA, USA). The extracted DNA was used to prepare libraries for sequencing with 2× Phanta Max Master Mix (VAZYME, China) polymerase. The V3–V4 variable region of bacterial 16S rDNA was amplified by PCR using the primers 338F: 5’-ACTCCTACGGGAGGCAGCAG-3’ and 806R: 5’-GGACTACHVGGGTWTCTAAT-3’. PCR enrichment was performed in a 50 µL reaction containing 30 ng of DNA template and fusion PCR primers. The PCR cycling conditions were as follows: 95°C for 3 min; 30 cycles of 95°C for 15 sec, 56°C for 15 sec, 72°C for 45 sec and final extension at 72°C for 5 min. PCR products were purified by DNA magnetic beads (BGI, LB00V60). The purified products were used to generate single-stranded library products through denaturation, followed by circularization to produce single-strand circular DNA molecules. Single-strand linear DNA was removed using digestion. The final single-strand circularized library is amplified with phi29 and rolling circle amplification (RCA) to generate DNA nano balls (DNB) carrying multiple copies of the initial single stranded library molecule. DNBs were loaded into the patterned nanoarray. Sequencing reads of PE300 bases length were generated with the DNBSEQ-G400 platform (BGI-Shenzhen, China), with a coverage of 100k reads (clean tags per sample). Raw sequencing data were processed using the QIIME2R-DADA2 workflow [41], an amplicon sequence variant (ASV)-based approach. Taxonomy was assigned using the SILVA database version 138.2. Microbiota diversity scores were determined using the Phyloseq and picante packages in R [42, 43].

### Primary outcomes

The primary outcomes were changes from baseline to the end of the six-month intervention in (1) CBF in our defined regions of interest (ROI), namely the dorsolateral prefrontal cortex (dlPFC) and hippocampus, (2) ROI-specific neural recruitment during a N-back working memory task, (3) N-back task accuracy (performance as assessed with d’), (4) plasma inflammation markers (hs-CRP, IL-6, and TNF-α), and (5) microbiota diversity scores (Shannon diversity, phylogenetic diversity, and Chao1 richness). We quantified CBF in ml/100g tissue/min, blood-oxygen-level-dependent (BOLD) activity as the measure of neurovascular coupling, and 2b-0b d’ performance scores as the measure for task accuracy.

### Secondary outcomes

Secondary outcomes included changes from baseline to the end of the six-month intervention in lifestyle-questionnaire measures of diet (DHD-index, MIND-NL), physical activity and sedentary behaviour (SQUASH, LASA), stress management and mindfulness (PSS, HADS, FFMQ), and sleep quality (PSQI). Additional secondary outcomes comprised of changes in anthropometric measurements, other brain health outcomes, cognitive functioning outcomes, cardiometabolic health indices, additional inflammation markers, intestinal integrity markers, and neurodegeneration-related biomarkers. The anthropometric measurements included BMI, waist-hip ratio, and blood pressure. The other brain health outcomes included dlPFC myo-inositol concentrations, and whole-brain GM and WM volumes. Cognitive functioning outcomes included all individual neuropsychological test outcomes, including the TMT, VFT, DSST, DST, RAVLT. The cardiometabolic health indices comprised glucose, insulin, cholesterol, HDL-cholesterol, LDL-cholesterol, and triglycerides levels. The inflammation markers comprised IFN-γ, IL-8, and IL-10 levels. The intestinal integrity markers measured were LPS, LBP, and zonulin. And last, the neurodegeneration-related biomarkers NfL, the Aβ42/Aβ40 ratio, and GFAP were considered.

### Data analysis

An estimated sample size of 104 participants was previously calculated to provide 80% power to detect an effect on the primary outcomes. In this calculation, we accounted for 15% data loss due to acquisition issues or drop-outs. Due to two last-minute cancellations, the final included sample size consisted of 102 participants.

Prior to analysis, we performed data cleaning and quality control assessments of all measures whilst blinded for intervention group allocation. Participants who clearly failed to perform the N-back task correctly, defined as having either a target accuracy <30% on the 0–back condition or a non-target accuracy <65% on any of the 0-, 1-, or 2-back conditions, were removed from further analysis (n=3 at baseline, n=2 at follow-up). fMRI data quality was subsequently assessed using the fMRIprep reports and motion metrics. Datasets with mean framewise displacement >0.5mm or maximum displacement >3mm underwent additional inspection. The first-level 2-back contrast map (thresholded at P=0.001 uncorrected) was visually examined for canonical fronto-parietal activation patterns associated with working memory. Datasets lacking any discernible fronto-parietal activity were excluded from analysis (n=5 at baseline, n=6 at follow-up). For pCASL, a number of datasets were excluded because of acquisition errors such as excessive movement (n=3 at baseline, n=2 at follow-up). For MRS, spectra at different processing stages (including pre-aligned, post-aligned, averaged, and fitted) were visually examined to assess overall quality. We evaluated lipid peak absence (1-2 ppm), sufficient water suppression (3-4 ppm), metabolite-of-interest peak presence (myo-inositol at 3.03 ppm), chemical shift drift absence (indicative of movement-related artifacts), and shimming effectivity as measures of sufficient data quality. Additionally, Osprey’s quality metrics, such as the creatine signal-to-noise ratio and FWHM were reviewed. Datasets that did not meet acceptable quality standards based on these metrics, were excluded from further analysis. Questionnaire data were manually checked for implausible entries (e.g., >7 days per week, more reported hours than possible wake time). Prior to analysis, assumption checks of data normality and linearity were performed with QQ- and scatterplots.

To test the effect of the lifestyle intervention on the primary and secondary outcomes, linear mixed-effect models with random effects for intercept (individuals) were performed. The intervention group and time were included as fixed effects, as well as the intervention term between intervention group and time to model the difference in change between baseline and follow-up as function of group allocation. False discovery rate (FDR) following the Benjamini-Hochberg (BH) procedure was separately applied for the primary outcomes and subgroups of secondary outcomes to adjust for multiple comparisons [44]. The analyses were conducted on an intention-to-treat basis. To explore the correlation between changes in individual delta (Δ, as T1 – T0) scores of (primary) brain and (primary) peripheral outcomes, we used Pearson correlation analyses. Changes in primary brain and peripheral outcomes were adjusted for their level at baseline using linear regression. Also, the CBF measures were adjusted for the pCASL sequence. Residuals from these models were extracted and used as adjusted measures of change. Pearson correlation coefficients were calculated between the residualized variables. The adjusted associations were visualized using scatterplots of the residuals with fitted linear regression lines and 95% confidence intervals. The P-values are not FDR-corrected due to the exploratory nature of the correlation analyses.

A sensitivity analyses was performed for the analyses focusing on CRP, in which participants with signs of acute infection, defined as CRP>22 mg/L, were excluded (n=2). CRP levels >22 mg/L indicate acute infection in overweight individuals or individuals with obesity [45, 46], which largely characterized our study population. All statistical analyses were performed using R software (version 4.3.2). Reported P-values throughout the manuscript are FDR-corrected, unless specified otherwise, and significance was set at PFDR<0.05 in two-tailed tests for all analyses.

## Results

### Characteristics of the study population

Between April 2022 and September 2023, 279 participants were screened for eligibility. Of these screened participants, 102 participants were randomised to either the active control group (n=49) or the intervention group (n=53) (**Figure 1**). Baseline outcome assessments started in May 2022 and the final follow-up assessment was completed in May 2024. In total, 16 participants fully discontinued their study participation and did not complete any follow-up assessments (active control group: n=10; intervention group: n=6), resulting in a total of 86 participants completing the six-month follow-up outcome assessments. Additionally, four participants in the intervention group discontinued the intervention but completed the follow-up assessments. Their data was therefore included in the intention-to-treat analyses.

**Figure 1.**
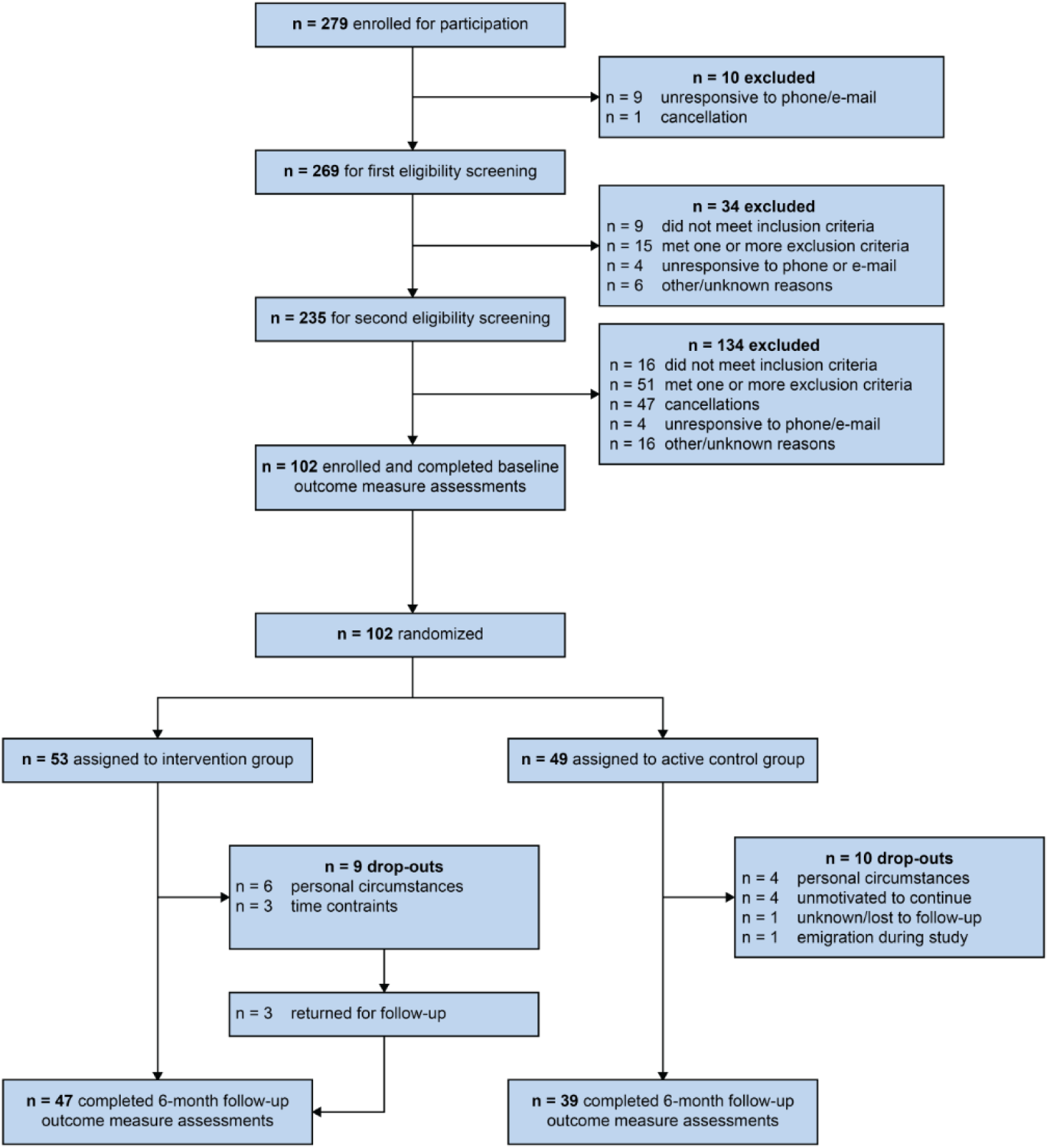
Flowchart of the study enrollment, inclusion, exclusion, randomisation, and outcome measure visits.

The active control and intervention group that completed baseline and six-month assessments had similar baseline characteristics (**Table 1**). The mean age was 66 (SD 4). Most participants were women (67%) and were high educated (57%). Most participants were relatively healthy at baseline in respect to mean DHD-index (100, SD 17) and mean total physical activity in minutes per week (6294, SD 3543).

**Table 1.** Baseline characteristics of the HELI study.

|  | **Intervention group**<br>**(n=53)** | **Active control group**<br>**(n=49)** |
| --- | --- | --- |
| **Demographics** |  |  |
| Age (years) | 66 (4) | 67 (4) |
| Sex |  |  |
| Female | 35 (66%) | 32 (65%) |
| Education level |  |  |
| Low | 5 (9%) | 5 (10%) |
| Medium | 19 (36%) | 16 (33%) |
| High | 29 (55%) | 28 (57%) |
| **Cardiovascular risk factors** |  |  |
| BMI $\geq 25$ | 42 (79%) | 34 (69%) |
| Physical inactivity | 27 (51%) | 30 (61%) |
| Hypertension | 32 (60%) | 26 (53%) |
| Untreated hypertension | 9 (17%) | 9 (18%) |
| Hypercholesterolemia | 31 (58%) | 26 (53%) |
| Diabetes type-2 | 10 (19%) | 10 (20%) |
| Mild cardiovascular disease | 2 (4%) | 9 (18%) |
| Risk factor score, median (IQR) | 3 (2.0, 3.0) | 3 (2.0, 4.0) |
| **Lifestyle factors** |  |  |
| Diet |  |  |
| DHD-index (/150) <sup>1</sup> | 100.38 (17.81) | 100.88 (16.42) |
| MIND-NL (/15) <sup>1</sup> | 8.39 (2.20) | 8.69 (1.86) |
| Physical activity |  |  |
| SQUASH (min/week) <sup>2</sup> | 7204.98 (3890.49) | 6986.15 (3530.70) |
| LASA-SBQ (min/day) <sup>2</sup> | 404.88 (191.51) | 376.91 (170.08) |
| Stress and mindfulness |  |  |
| PSS (/40) <sup>3</sup> | 12.30 (4.53) | 12.53 (3.85) |
| HADS (/42) <sup>3</sup> | 7.51 (5.43) | 7.31 (4.84) |
| FFMQ (/195) <sup>3</sup> | 87.41 (9.90) | 87.28 (7.72) |
| Sleep |  |  |
| PSQI (/21) <sup>4</sup> | 6.83 (3.77) | 5.45 (3.16) |

Data are presented as mean ( ± SD) for continuous variables, and *n* (%) for categorical variables, unless specified otherwise.
<sup>1</sup> Higher scores indicate higher adherence to recommendations.
<sup>2</sup> Higher scores indicate higher physical/sedentary behaviour for the SQUASH or LASA-SBQ, respectively.
<sup>3</sup> Higher scored indicate greater perceived stress (PSS), anxiety/depression symptoms (HADS), and mindfulness traits (FFMQ).
<sup>4</sup> Higher scores indicate worse sleep quality.
Abbreviations: BMI, body mass index; DHD, Dutch healthy diet; FFMQ, Five Facet Mindfulness Questionnaire; HADS, Hospital and Anxiety Depression Scale; IQR, interquartile range; PSS, perceived stress scale; PSQI, Pittsburgh Sleep Quality Index; SBQ, sedentary behaviour questionnaire.

### Lifestyle intervention domains

When examining within-group effects for the lifestyle factors, we found that the DHD-index, MIND-diet, and sleep quality significantly increased during the intervention in the intervention group (PFDR=<0.0001, 0.0001, and 0.0003, respectively) but not in the active control group (**Supplementary File 2**). The HADS score decreased in the intervention group, but this was not significant after FDR correction (Punadjusted=0.03, PFDR=0.06). With regard to between-group effects, the intervention resulted in improved MIND-NL diet and sleep scores in the intervention group compared to the active control group (Beta 1.67, 95% CI 0.61;2.73, PFDR=0.01, and Beta −1.80, 95% CI −2.79;-0.81, PFDR=0.005, respectively) (**Table 2**). In the other three lifestyle intervention domains (physical activity, stress management and mindfulness, and cognitive training), there were no significant within-group or between-group differences (**Table 2**, **Supplementary File 2**).

**Table 2.** Results of intervention on the different lifestyle intervention domains of the HELI study.

|  | Intervention group |  |  |  | Active control group |  |  |  |  |  |  |
| --- | --- | --- | --- | --- | --- | --- | --- | --- | --- | --- | --- |
|  | Baseline |  | Six-month |  | Baseline |  | Six-month |  | Between-group difference |  |  |
|  | n | Mean ±SD | n | Mean ±SD | n | Mean ±SD | n | Mean ±SD | Mean change (95% CI) <sup>1</sup> | Unadjusted P-values | FDR adjusted P-values |
| **Lifestyle questionnaires** |  |  |  |  |  |  |  |  |  |  |  |
| DHD-index (/150) | 53 | 100 ±18 | 47 | 112 ±19 | 49 | 101 ±16 | 39 | 106 ±17 | 6.69 (0.20;13.19) | 0.05 | 0.13 |
| MIND-NL (/15) | 53 | 8.4 ±2.2 | 46 | 10.5 ±2.2 | 47 | 8.7 ±1.9 | 36 | 9.1 ±1.7 | **1.67 (0.61;2.73)** | **0.003** | **0.01** |
| SQUASH (min/week) | 53 | 6300 ±3710 | 47 | 7400 ±4660 | 48 | 6290 ±3390 | 36 | 6990 ±4810 | 297.54 (-1330.91;1930.56) | 0.72 | 0.72 |
| LASA-SBQ (min/day) | 53 | 462 ±219 | 47 | 502 ±201 | 49 | 493 ±205 | 37 | 479 ±249 | 51.04 (-51.19;153.63) | 0.33 | 0.38 |
| PSS (/40) | 53 | 15.1 ±5.0 | 47 | 14.9 ±5.6 | 49 | 15.1 ±4.8 | 38 | 13.9 ±5.4 | 1.13 (-0.53;2.79) | 0.19 | 0.30 |
| HADS (/42) | 53 | 7.5 ±5.4 | 47 | 6.2 ±4.7 | 49 | 7.3 ±4.8 | 37 | 7.0 ±4.9 | -0.99 (-2.75; 0.76) | 0.27 | 0.36 |
| FFMQ (/195) | 53 | 87.3 ±7.8 | 48 | 86.7 ±8.5 | 49 | 87.4 ±10.0 | 38 | 89.4 ±9.7 | -2.50 (-5.29;0.30) | 0.08 | 0.16 |
| PSQI (/21) | 53 | 6.7 ±3.6 | 47 | 5.3 ±3.0 | 49 | 5.4 ±3.1 | 37 | 5.6 ±3.3 | **-1.80 (-2.79;-0.81)** | **<0.001** | **0.005** |
| **Cognitive assessment** |  |  |  |  |  |  |  |  |  |  |  |
| TMT (TMT-B/TMT-A) | 53 | 2.31 ±0.82 | 47 | 2.20 ±0.63 | 49 | 2.17 | 39 | 2.20 | -0.13 (-0.43;0.17) | 0.39 | 0.93 |
| ratio score)<sup>2</sup> |  |  |  |  |  | ±0.55 |  | ±0.70 |  |  |  |
| VFT (total score) | 53 | 26.0 ±6.06 | 47 | 26.1 ±5.31 | 49 | 26.0<br>±4.89 | 39 | 26.6<br>±5.69 | -0.20 (-2.05;1.65) | 0.84 | 0.93 |
| DSST (total score) | 53 | 52.6 ±9.70 | 47 | 54.9 ±10.0 | 49 | 51.3<br>±10.6 | 39 | 53.1<br>±10.3 | -0.11 (-2.49;2.28) | 0.93 | 0.93 |
| RAVLT delayed recall (/15) | 53 | 7.77 ±3.24 | 47 | 10.3 ±3.21 | 49 | 7.51<br>±3.12 | 39 | 9.59<br>±3.36 | 0.18 (-0.96;1.32) | 0.76 | 0.93 |
| WAIS digit span (/28) | 53 | 15.5 ±3.69 | 47 | 15.4 ±3.77 | 49 | 14.9<br>±3.10 | 39 | 15.3<br>±3.47 | -0.95 (-1.94;0.05) | 0.06 | 0.30 |

Numbers in between brackets behind the tests and questionnaires reflect the maximum score.
<sup>1</sup> Calculated using linear mixed-effect models with random effects of intercept (individuals) and group and time as fixed effects, as well as the interaction term between intervention group and time. The results of the interaction term between intervention group and time are shown here.
<sup>2</sup> To account for TMT-A completion time, a ratio score was calculated by dividing the TMT-B scores with the TMT-A scores (TMT-B/TMT-A)
Abbreviations: DHD, Dutch healthy diet; FFMQ, Five Facet Mindfulness Questionnaire; HADS, Hospital and Anxiety Depression Scale; LASA, Sedentary Behaviour Questionnaire; PSS, Perceived Stress Scale; PSQI, Pittsburgh Sleep Quality Index; SQUASH, Short Questionnaire to Assess Health-enhancing physical activity.

For cognition, the DSST and RAVLT significantly increased in both the intervention (PFDR=0.03, and 0.0001, respectively) and the active control group (PFDR=0.03, and 0.0001, respectively) (**Supplementary File 2**), but the improvement in these or other cognitive outcomes did not significantly differ between the groups (**Table 2**).

### Primary outcomes

In the active control group, CBF increased in the dlPFC (Punadjusted<0.001, PFDR=0.009) and hippocampus (Punadjusted =0.02), but hippocampal CBF did not remain significant after FDR correction (PFDR =0.11). There were no other significant within-group effects for the primary brain outcomes (**Supplementary File 3**). When examining differences between the groups, we found that the intervention resulted in decreased CBF in the hippocampus in the intervention group compared to the active control group (Beta −6.62, 95% CI −11.23;-2.07, Punadjusted=0.006), however, this was not significant after applying FDR correction (PFDR=0.07). There were no other between-group differences for the primary brain outcomes (**Table 3**).

**Table 3.**
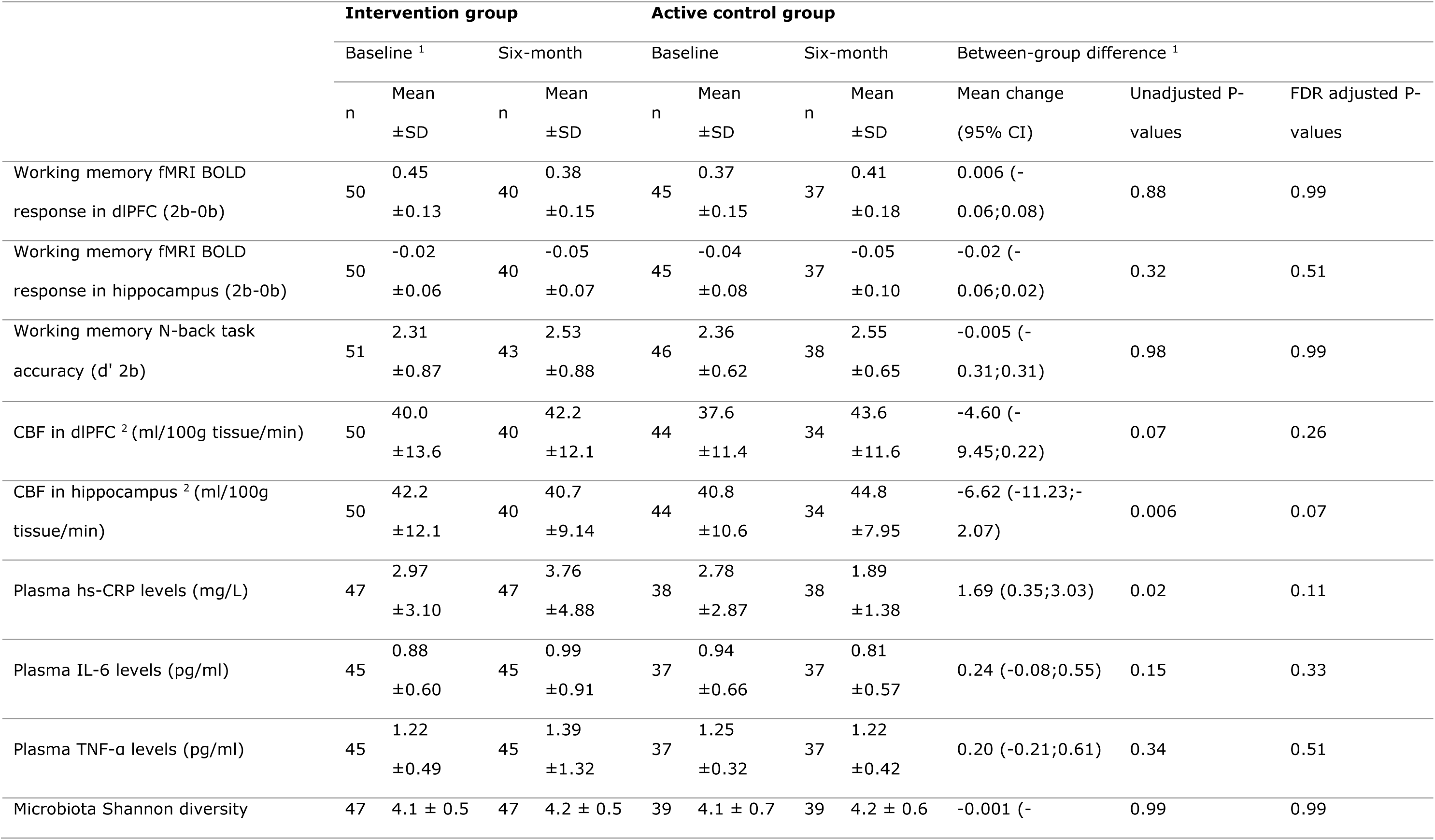
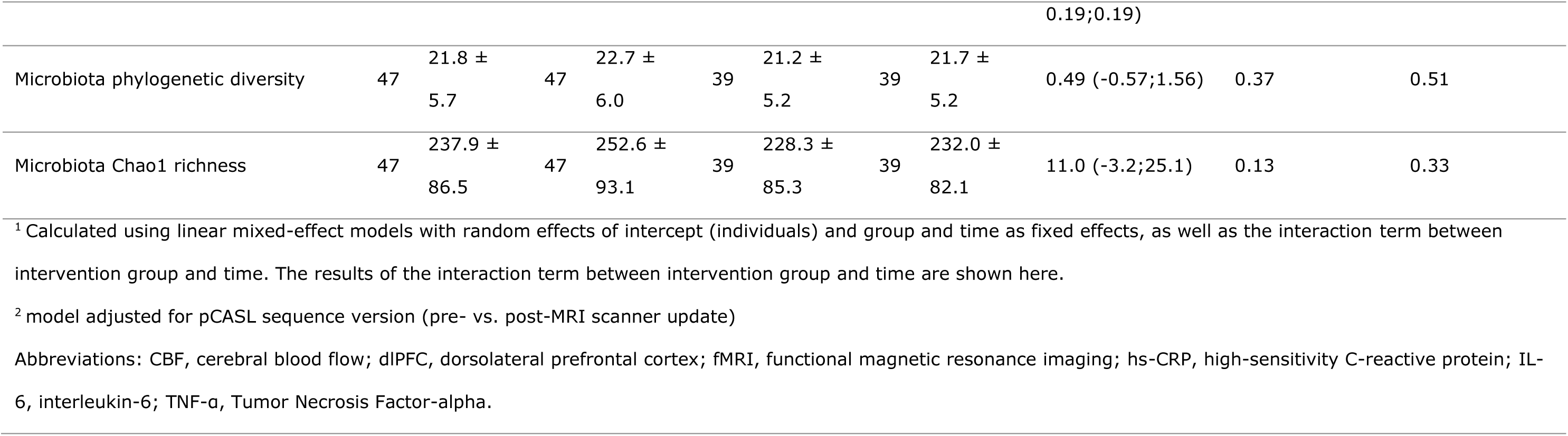
Results of the primary outcomes of the HELI study.

There were no within-group differences for the primary inflammation outcomes (**Supplementary File 3**). The CRP levels increased over time in the intervention group compared to the active control group, however, the FDR-corrected P-value was not significant (Beta 1.69, 95% CI 0.35;3.03, Punadjusted=0.02, PFDR=0.11) (**Table 3**). In the sensitivity analysis excluding participants with suspected acute infections (n=2), the between-group differences remained similar (Beta 1.16, 95% CI 0.12;2.19, Punadjusted=0.03, PFDR=0.17). There were no other between-group differences for the primary inflammation outcomes.

Examining within-group differences in the microbiota outcomes revealed that the microbiota phylogenetic diversity score increased in the intervention group, but not significant after FDR correction (Punadjusted=0.02, PFDR=0.11). The Chao1 richness index significantly increased in the intervention group (PFDR=0.03) (**Supplementary File 3**). There were no between-group differences for the primary microbiota diversity outcomes (**Table 3**).

In the total sample, irrespective of intervention group allocation, an increase in individual CRP levels over time was correlated with a reduction in CBF between baseline and six-month follow-up in the hippocampus (Punadjusted=0.03) (**Figure 3A**). However, this correlation disappeared after excluding participants with acute infections (Punadjusted=0.09) (**Supplementary File 4**). There were no statistically significant correlations between changes in the other primary brain and primary peripheral outcomes (**Figure 2-6**).

**Figure 2.**
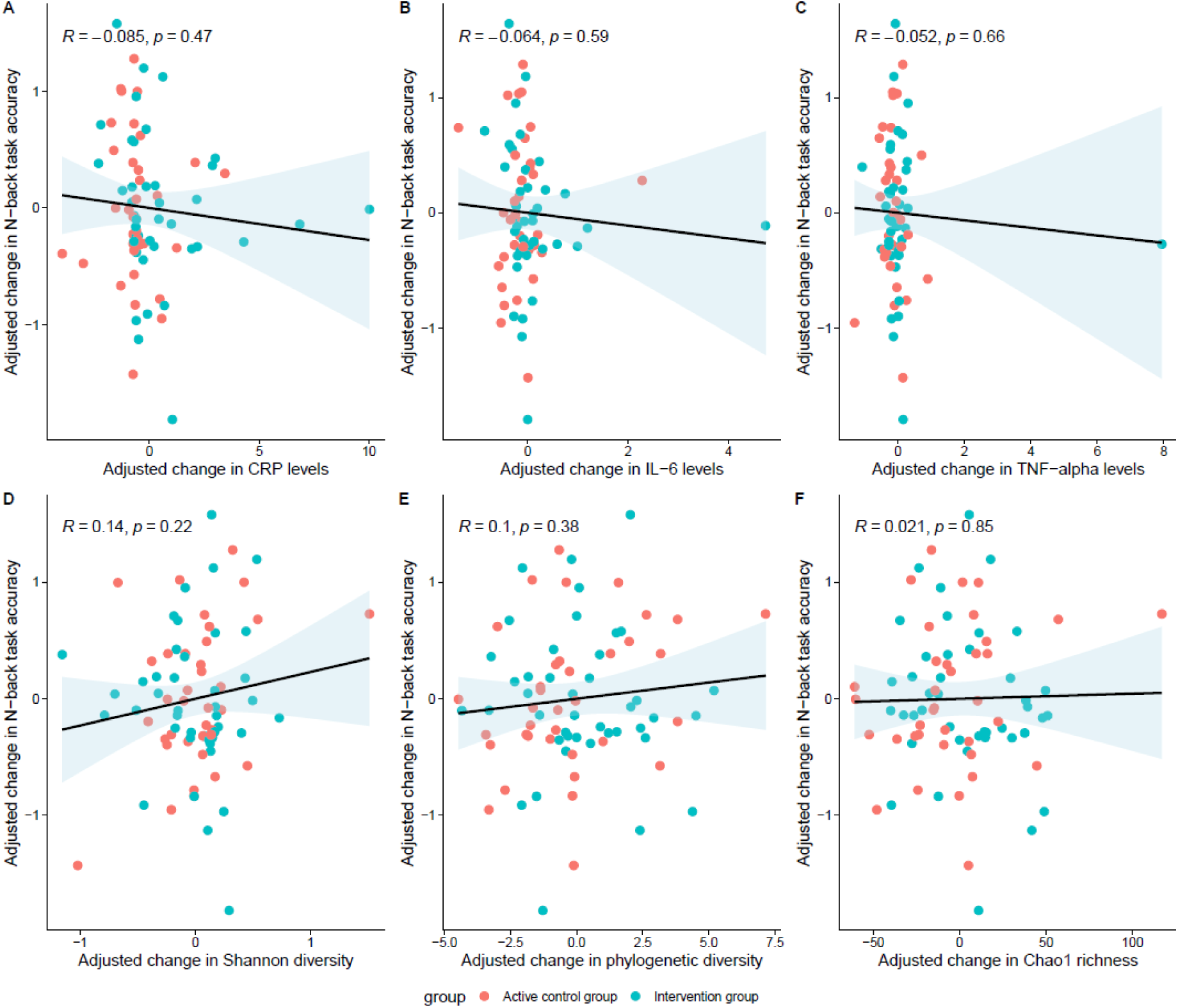
The correlation between change in N-back task accuracy and change in primary peripheral outcomes (A: n=75, B: n=74, C: n=74, D: n=78, E: n=78, F: n=78). Changes in N-back task accuracy and primary peripheral outcomes were adjusted for their baseline levels before calculation of the Pearson correlation. Data points are colored by intervention group. The solid line indicates the fitted regression line and the shaded area represents the 95% confidence interval. P-values are not FDR corrected.

**Figure 3.**
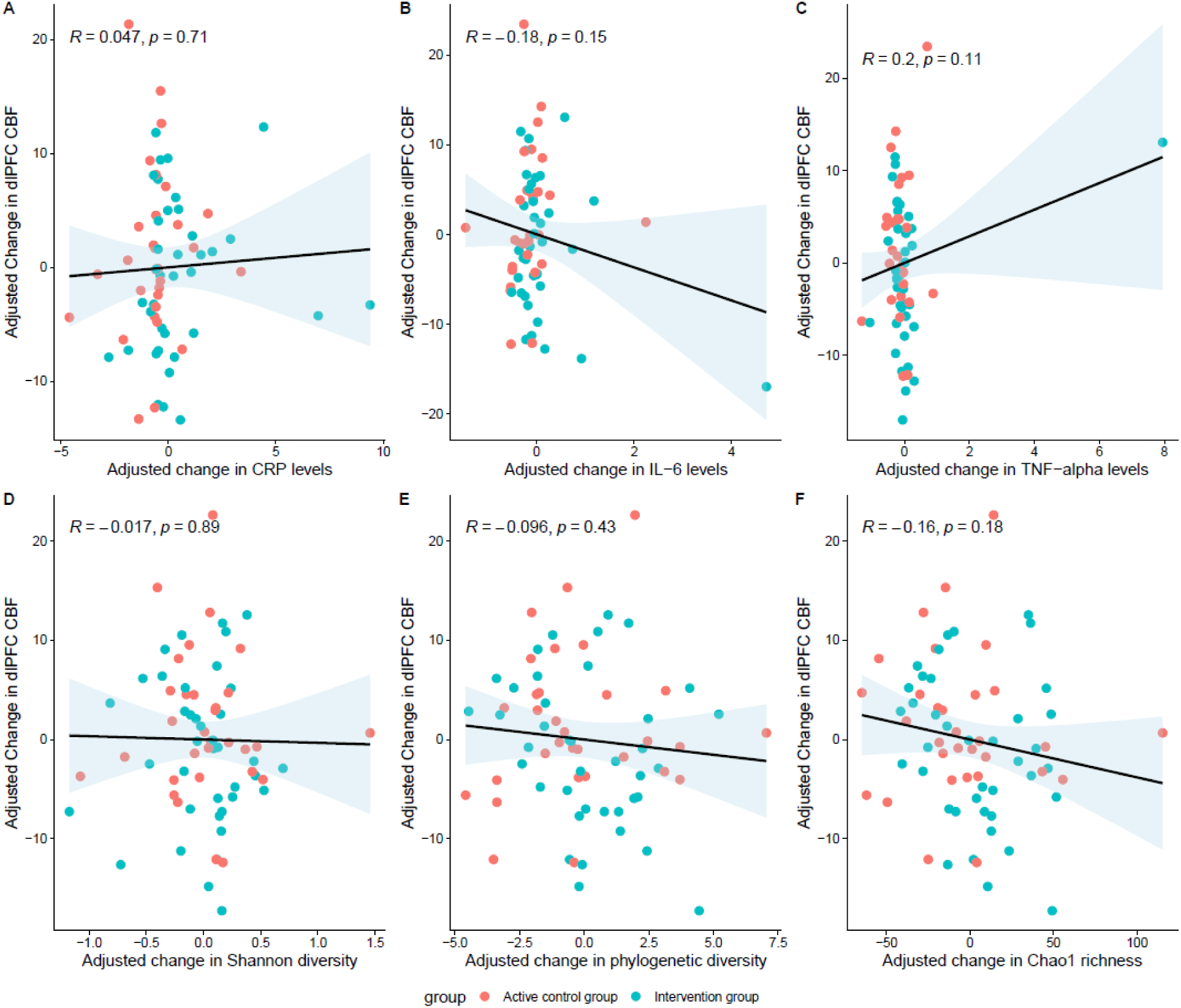
The correlation between change in CBF in dlPFC and change in primary peripheral outcomes (A: n=67, B: n=66, C: n=66, D: n=70, E: n=70, F: n=70). Changes in dlPFC CBF and primary peripheral outcomes were adjusted for their baseline levels before calculation of the Pearson correlation. Also, the CBF measures were adjusted for the pCASL sequence. Data points are colored by intervention group. The solid line indicates the fitted regression line and the shaded area represents the 95% confidence interval. P-values are not FDR corrected.

**Figure 4.**
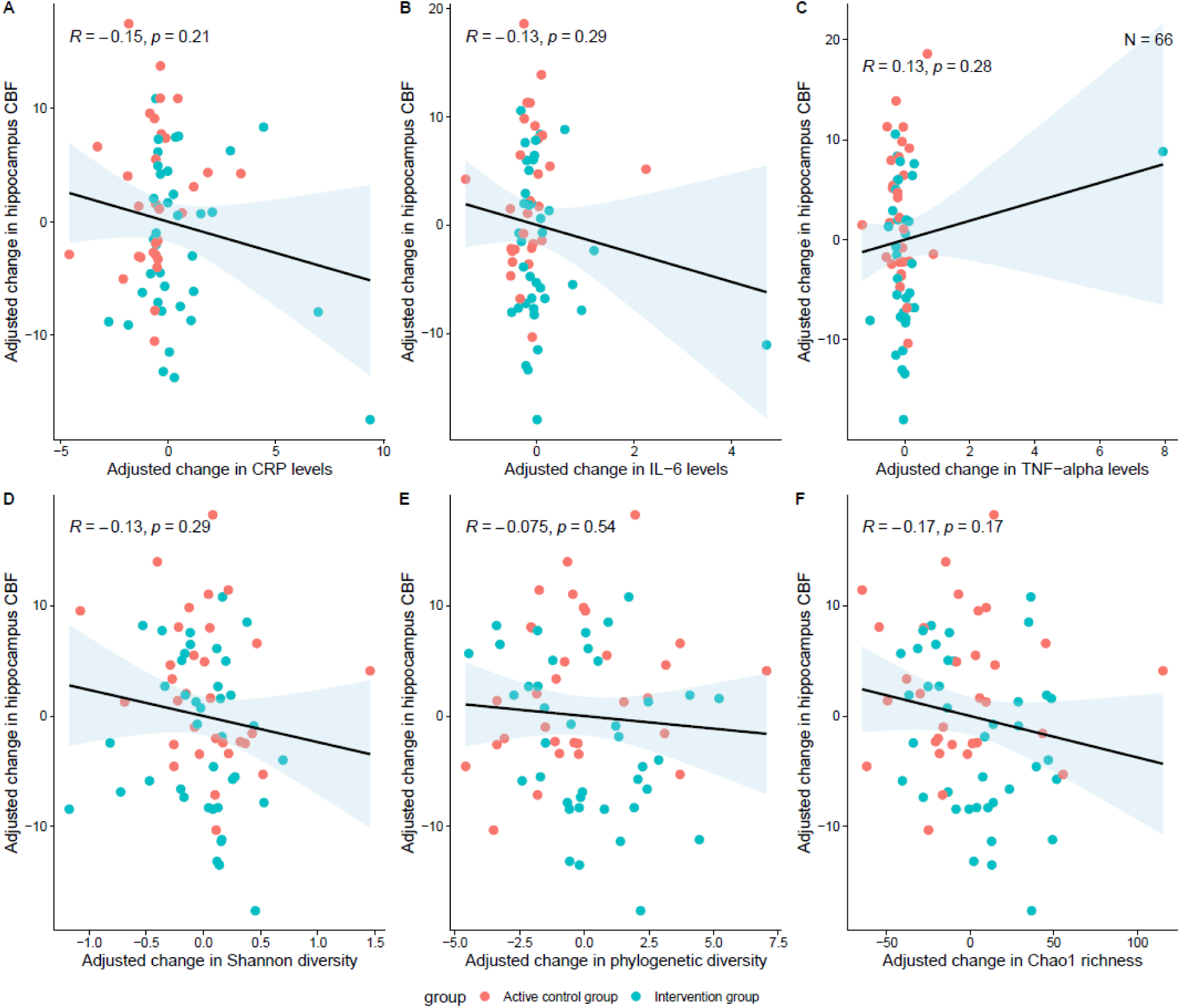
The correlation between change in CBF in hippocampus and change in primary peripheral outcomes (A: n=67, B: n=66, C: n=66, D: n=70, E: n=70, F: n=70). Correlation coefficient and P-values are calculated using Pearson correlation. Changes in hippocampus CBF and primary peripheral outcomes were adjusted for their baseline levels before calculation of the Pearson correlation. Also, the CBF measures were adjusted for the pCASL sequence. Data points are colored by intervention group. The solid line indicates the fitted regression line and the shaded area represents the 95% confidence interval. P-values are not FDR corrected.

**Figure 5.**
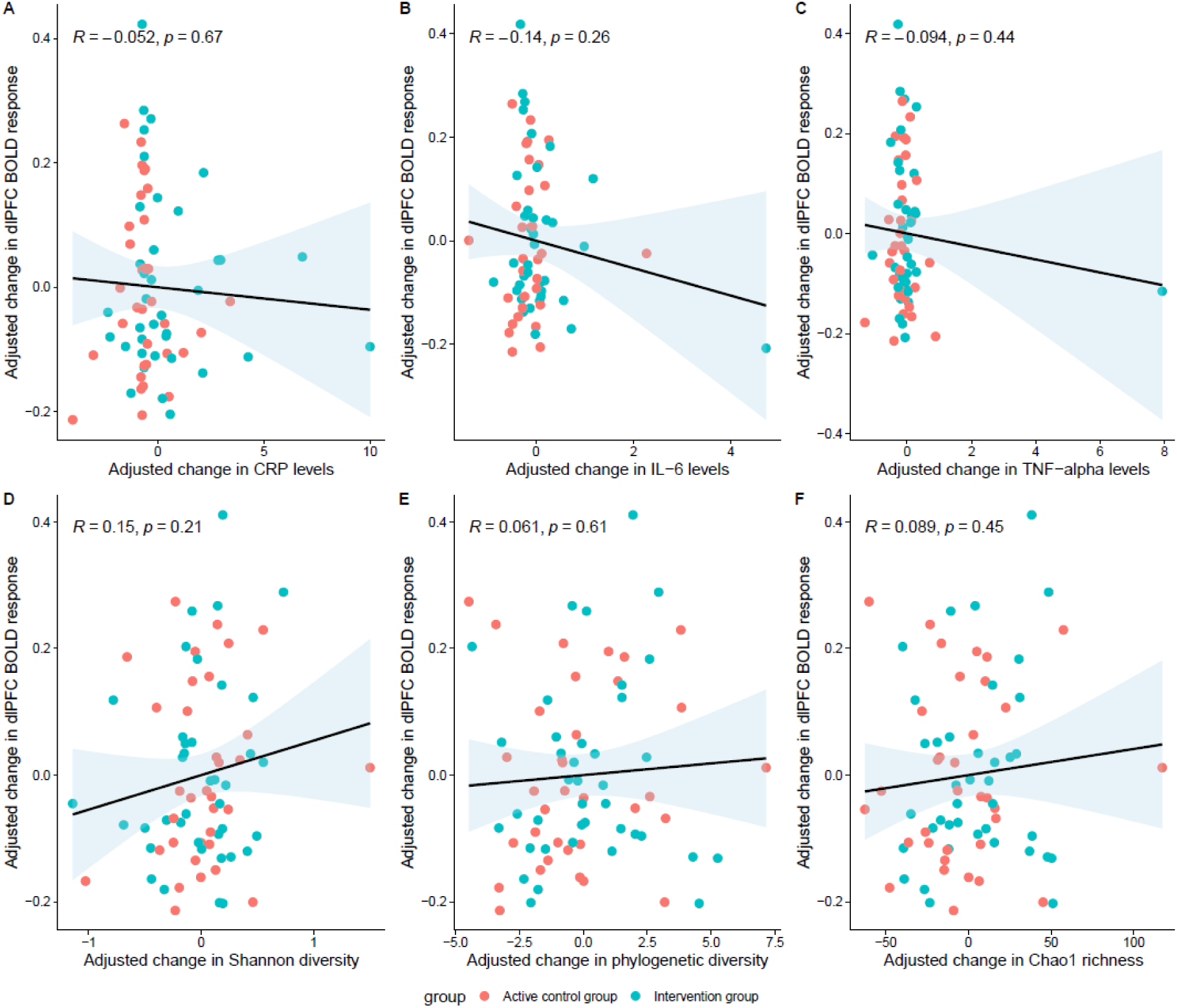
The correlation between change in fMRI BOLD responses in dlPFC and change in primary peripheral outcomes (A: n=70, B: n=69, C: n=69, D: n=73, E: n=73, F: n=73). Changes in dlPFC BOLD response and primary peripheral outcomes were adjusted for their baseline levels before calculation of the Pearson correlation. Data points are colored by intervention group. The solid line indicates the fitted regression line and the shaded area represents the 95% confidence interval. P-values are not FDR corrected.

**Figure 6.**
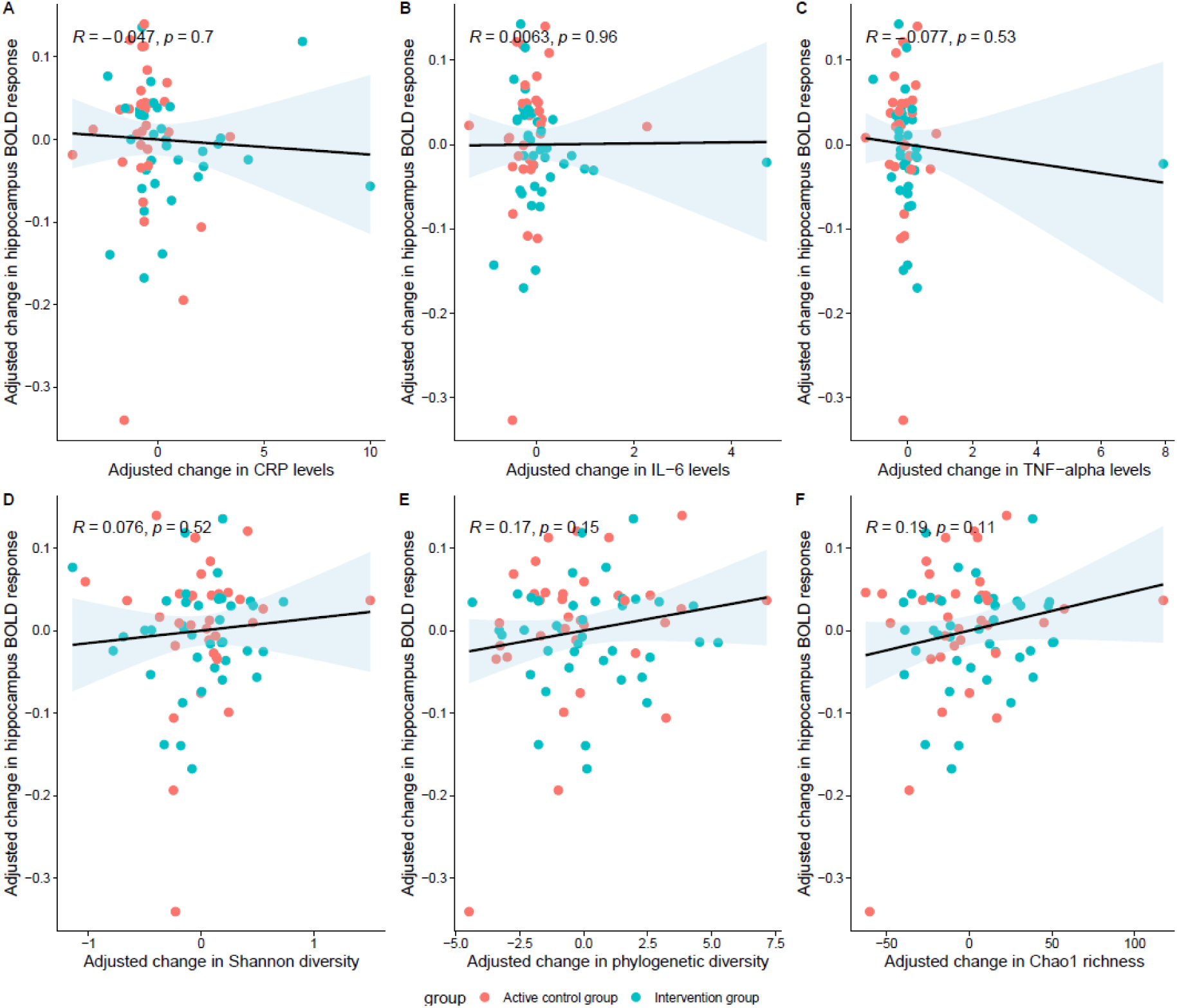
The correlation between change in fMRI BOLD responses in hippocampus and change in primary peripheral outcomes (A: n=70, B: n=69, C: n=69, D: n=73, E: n=73, F: n=73). Changes in hippocampus BOLD response and primary peripheral outcomes were adjusted for their baseline levels before calculation of the Pearson correlation. Data points are colored by intervention group. The solid line indicates the fitted regression line and the shaded area represents the 95% confidence interval. P-values are not FDR corrected.

### Secondary outcomes

Within groups, significant reductions in BMI, waist-hip ratio, and blood pressure were observed in the intervention group (PFDR=0.02, 0.008, and 0.0006, respectively) (**Supplementary File 5**; however, there were no significant between-group differences for BMI, waist-hip ratio, and blood pressure (**Table 4**). For cardiometabolic markers measured in blood, levels of cholesterol, LDL-cholesterol, and triglycerides significantly decreased during the intervention period in the intervention (PFDR=0.01, 0.01, and 0.01, respectively). The triglycerides levels decreased in the intervention compared to the active control group over time, but this was not significant after FDR correction (Beta −0.22, 95% CI −0.40; −0.04, Punadjusted=0.02, PFDR=0.12). For the other cardiometabolic markers, there were no between-group differences. The levels of NfL and GFAP, biomarkers of neurodegeneration, significantly increased over time in the active control group (PFDR=0.02 and 0.002, respectively). No between-group differences were observed for the biomarkers of neurodegeneration. For the other secondary outcomes, there were no within-group or between-group differences (**Table 4**, **Supplementary File 5**).

**Table 4.** Results of the secondary outcomes of the HELI study.

|  | Intervention group |  |  |  | Active control group |  |  |  | Between-group difference |  |  |
| --- | --- | --- | --- | --- | --- | --- | --- | --- | --- | --- | --- |
|  | Baseline |  | Six-month |  | Baseline |  | Six-month |  |  |  |  |
|  | n | Mean ±SD | n | Mean ±SD | n | Mean ±SD | n | Mean ±SD | Mean change (95% CI)<sup>1</sup> | Unadjusted P-values | FDR adjusted P-values<sup>2</sup> |
| Anthropometric measurements |  |  |  |  |  |  |  |  |  |  |  |
| BMI (kg/m²) | 53 | 31.0 ±5.9 | 47 | 30.3 ± 5.7 | 49 | 28.9 ±4.7 | 39 | 28.4 ±4.7 | -0.51 (-1.12;1.10) | 0.11 | 0.12 |
| Waist-hip ratio | 53 | 0.97 ±0.1 | 47 | 0.93 ±0.1 | 49 | 0.94 ±0.1 | 39 | 0.93 ±0.1 | -0.03 (-0.07;0.01) | 0.12 | 0.12 |
| Blood pressure (mmHg) | 52 | 110 ±14 | 46 | 105 ±11 | 49 | 107 ±13 | 39 | 104 ±12 | -3.10 (-6.90;0.72) | 0.12 | 0.12 |
| Cardiometabolic profile |  |  |  |  |  |  |  |  |  |  |  |
| Glucose (mmol/L) | 53 | 6.7 ±2.5 | 46 | 6.3 ±1.7 | 48 | 6.3 ±1.3 | 36 | 6.2 ±1.6 | -0.38 (-0.94;0.19) | 0.19 | 0.29 |
| Insulin (mmol/L) | 49 | 24.7 ±58.2 | 46 | 24.3 ±60.1 | 45 | 13.6 ±8.8 | 36 | 12.9 ±8.5 | 0.55 (-1.67;2.80) | 0.63 | 0.76 |
| Cholesterol (mmol/L) | 53 | 5.5 ±1.0 | 46 | 5.2 ±1.0 | 48 | 5.7 ±1.3 | 37 | 5.7 ±1.3 | -0.25 (-0.58;0.08) | 0.14 | 0.28 |
| HDL-cholesterol (mmol/L) | 53 | 1.5 ±0.4 | 46 | 1.5 ±0.4 | 48 | 1.4 ±0.4 | 37 | 1.5 ±0.4 | -0.01 (-0.09;0.07) | 0.78 | 0.78 |
| LDL-cholesterol | 53 | 3.3 ±0.9 | 46 | 3.0 ±0.8 | 48 | 3.5 ±1.1 | 37 | 3.5 ±1.0 | -0.23 (-0.51;0.05) | 0.11 | 0.28 |
| (mmol/L) |  |  |  |  |  |  |  |  |  |  |  |
| Triglycerides | 53 | 1.7 ±0.7 | 46 | 1.6 ±0.7 | 48 | 1.6 ±0.8 | 37 | 1.6 ±0.7 | -0.22 (-0.40;-0.04) | 0.02 | 0.12 |
| (mmol/L) |  |  |  |  |  |  |  |  |  |  |  |
| **Neuroimaging** |  |  |  |  |  |  |  |  |  |  |  |
| Hippocampal volume<br>³ (mm³) | 53 | 7870 ±765 | 45 | 7830 ±702 | 48 | 7760<br>±777 | 38 | 7720<br>±763 | -17.7 (-87.42;<br>51.95) | 0.81 | 0.81 |
| dIPFC volume ³<br>(mm³) | 53 | 24500<br>±2830 | 45 | 24600 ±3030 | 48 | 23700<br>±2930 | 38 | 23600<br>±2580 | -11.24 (-455.42;<br>432.95) | 0.80 | 0.81 |
| Brain myo-inositol<br>levels (mol/kg) | 52 | 10.3 ±3.4 | 45 | 9.8 ±1.3 | 48 | 10.4<br>±1.9 | 36 | 10.6<br>±2.5 | -0.24 (-1.21; 0.72) | 0.55 | 0.81 |
| **Inflammation profile** |  |  |  |  |  |  |  |  |  |  |  |
| IFN-γ (pg/mL) | 45 | 4.49 ±2.48 | 45 | 9.44 ±28.9 | 37 | 6.17<br>±7.59 | 37 | 5.51<br>±5.31 | 5.61 (-4.08;15.30) | 0.26 | 0.50 |
| IL-8 (pg/mL) | 45 | 4.57 ±1.30 | 45 | 4.54 ±1.56 | 37 | 4.70<br>±1.18 | 37 | 4.75<br>±1.69 | -0.07 (-0.64;0.51) | 0.82 | 0.82 |
| IL-10 (pg/mL) | 45 | 0.31 ±0.79 | 45 | 0.69 ±3.27 | 37 | 0.28<br>±0.19 | 37 | 0.25<br>±0.15 | 0.40 (-0.40;1.20) | 0.33 | 0.50 |
| **Biomarkers of intestinal integrity** |  |  |  |  |  |  |  |  |  |  |  |
| LBP (µg/ml) | 39 | 13.7 ±4.1 | 38 | 13.9 ±4.4 | 36 | 12.8<br>±3.7 | 35 | 12.8<br>±3.5 | 1.12 (-0.19;2.42) | 0.10 | 0.30 |
| LPS ( EU/ml) | 39 | 4.3 ±13.7 | 38 | 26.2 ±109.0 | 36 | 17.8<br>±65.9 | 35 | 19.1<br>±59.7 | 16.07 (-<br>24.43;56.57) | 0.44 | 0.44 |
| Zonulin (ng/ml) | 39 | 40.3 ±16.7 | 38 | 39.3 ±17.0 | 36 | 37.2<br>±17.0 | 35 | 36.0<br>±15.1 | 2.01 (-1.55;5.56) | 0.27 | 0.41 |
| **Biomarkers of neurodegeneration** |  |  |  |  |  |  |  |  |  |  |  |
| NfL (pg/mL) | 47 | 15.0 ±6.1 | 46 | 15.0 ±5.6 | 38 | 13.1<br>±5.5 | 36 | 14.2<br>±6.2 | -1.09 (-2.18;0.001) | 0.05 | 0.15 |
| Aβ42/40 ratio | 47 | 0.057<br>±0.011 | 46 | 0.057 ±0.012 | 38 | 0.058<br>±0.011 | 36 | 0.058<br>±0.011 | -0.001 (-<br>0.003;0.001) | 0.28 | 0.28 |
| GFAP (pg/mL) | 47 | 77.0 ±24.7 | 46 | 80.0 ±29.5 | 38 | 71.9<br>±21.4 | 36 | 79.1<br>±24.7 | -3.77 (-8.92;1.38) | 0.16 | 0.24 |
| **Other** |  |  |  |  |  |  |  |  |  |  |  |
| White blood cell<br>count (counts×10<sup>9</sup>/L) | 53 | 6.2 ±1.9 | 47 | 6.5 ±2.5 | 49 | 6.3 ±1.8 | 38 | 5.2 ±1.8 | 0.45 (-0.35;1.26) | 0.27 | - |
| <sup>1</sup> Calculated using linear mixed-effect models with random effects of intercept (individuals) and group and time as fixed effects, as well as the interaction term between intervention group and time. The results of the interaction term between intervention group and time are shown here.<br><sup>2</sup> FDR-corrected per subgroup.<br><sup>3</sup> Model adjusted for total brain volume. |  |  |  |  |  |  |  |  |  |  |  |

## Discussion

With this randomised controlled trial, we aimed to investigate brain, immune, and gut factors that could underlie the effects of multidomain lifestyle interventions on cognitive functioning in older individuals as they were reported in previous trials. Together, the results show that the six-month multidomain lifestyle intervention was able to improve diet and sleep quality scores. Also, within the intervention group, favourable reductions in lifestyle-modifiable cardiovascular risk factors, such as BMI and blood pressure, were observed, but these were not significantly different from changes in the active control group. Moreover, the lifestyle intervention did not have an acute effect on various brain, metabolic, inflammatory, and gut health measures included in this study.

The current intervention consisted of five different lifestyle domains: diet, physical activity, stress management and mindfulness, cognitive training, and sleep. We only observed within-group and between-group intervention effects on diet and sleep, but not for physical activity, stress management and mindfulness, and cognitive training. Moreover, both the intervention and active control group showed significant improvements in cognition over time, potentially reflecting learning effects. These results suggest that the intervention was able to provide meaningful impact on two out of the five targeted lifestyle domains (diet and sleep), but was unable to capture significant effects on the other three target lifestyle domains (physical activity, stress management and mindfulness, cognitive training).

We intentionally chose not to select participants based on how healthy their lifestyle was at baseline, besides physical activity, arguing that every participant could benefit from making changes to their lifestyle regardless of their starting point, i.e., either by maintaining already high levels or by improving their level. This resulted in a selection of participants who complied relatively well with lifestyle guidelines/advice at baseline compared to the general Dutch population. For example, the mean DHD-index of the HELI participants at baseline was 100 (out of 150) compared to 75-81 (out of 160) in the general Dutch population [47]. Also, improvements in the active control group may contribute to reduced intervention effects. For example, independent of group allocation, we found some careful suggestions that individual changes over time in CRP levels may be correlated with changes over time in CBF in the hippocampus and dlPFC. In addition, the information received by the active control group in the form of leaflets on general lifestyle-related health may be already sufficient information or an incentive needed for individuals to make lifestyle adjustments, so that personal lifestyle advice and group sessions may become unnecessary. However, it is important to note that the participants in the HELI study were not only relatively highly educated and healthy at baseline but also decided to participate in a trial on lifestyle, implying their willingness to make lifestyle changes. Future research is warranted to investigate the effect of lifestyle changes on brain function and metabolic health in individuals who do not yet adhere to dietary and lifestyle recommendations.

The six-month lifestyle intervention did not have any effects on our primary neuroimaging measures of brain functioning (task-based BOLD response and CBF in dlPFC and hippocampus) or secondary measures of ROI brain volumetrics and MRS. Previous studies, targeting only one or two lifestyle domains, have shown beneficial effects of improved diet, physical activity, and cognitive training on neural recruitment [48] and CBF [49–51]. Also, in the Multidomain Alzheimer Prevention Trial (MAPT), the lifestyle intervention group showed decreased deformation of periventricular areas and temporal lobes and decreased whole-brain atrophy compared to placebo [9]. However, within the Finnish Geriatric Intervention Study to Prevent Cognitive Impairment and Disability (FINGER) study, no structural (i.e., anatomical) differences were found between intervention groups [52], similar to our findings.

It is possible that our intervention duration of six months was not sufficiently long enough to capture the true associations between changes in lifestyle patterns, reduced cardiovascular risk, and subsequent changes in structural and functional brain mechanisms, as longer multidomain lifestyle intervention trials such as the German Agewell.de trial have shown effects on whole-brain GM CBF over four years [12]. Moreover, in the Japan-Multimodal Intervention Trial (JMINT) trial, the intervention group showed greater reductions compared to the control group in anthropometric measurements, including decreased body fat percentage and free-fat mass after 6 and 18 months. However, body free fat percentage and free-fat mass were not measured in the current study. Interestingly, changes in BMI and SBP were only observed after 18 months, whereas no significant effects were observed at the 6-month follow-up [15]. Additionally, the FINGER trial reported that the intervention group showed greater improvements compared to the control group in BMI reduction, intake of fish and vegetables, and physical activity after two years [7]. It is important to note, however, that direct comparisons between HELI and AgeWell.de, JMINT, and the FINGER trial are challenging due to the differences in assessment of lifestyle and vascular factors (e.g., HELI assessed physical activity using the more comprehensive SQUASH questionnaire, while the FINGER trial used a cut-off of two physical activities per week as a measure of success) and study duration. Also, sample sizes of these trials are substantially larger compared to our study (n=461 in AgeWell.de, n=406 in JMINT, and n=1260 in FINGER trial). Nevertheless, unimodal intervention studies with smaller sample sizes (n=26-41) focusing primarily on diet or physical exercise have reported significant increases in brain activation within (pre)frontal ROIs, using intervention durations between 12 weeks and 6 months [53, 54]. These findings suggest that perhaps unimodal programs may be particularly influential in modulating fMRI-based brain activation measures, and the inclusion of additional domains may have diluted the specific effects of certain specific domains. Moreover, our aim was to explore potential underlying brain and peripheral mechanisms of multidomain lifestyle interventions that potentially drive reported cognitive benefits of multidomain lifestyle interventions. To this end, we assessed a comprehensive range of cognitive, neurobiological, and peripheral outcomes measures. Although this approach provided a broad characterization of potential intervention-related effects, the large number of outcomes necessitated extensive correction for multiple testing, reducing our ability to detect modest intervention effects.

The HELI study is the first to investigate an extensive array of brain and peripheral health outcome measures to assess the effects underlying a multidomain lifestyle intervention in an older population at risk of cognitive decline. A major strength of our study is the use of an extensive neuroimaging protocol, which includes multiple functional brain measures potentially associated with cognitive improvement reported in other multidomain lifestyle intervention studies such as the FINGER and US-POINTER trials. Most previous work on the mechanisms underlying cognitive decline has focused on either healthy older adults or clinical populations, such as mild cognitive impairment (MCI) and early Alzheimer’s disease (AD). Populations of people with MCI specifically are often used as an intermediate stage between healthy and pathological aging [55], but the mechanisms involved may be quite different as cognitive decline is already in a progressed stage. In contrast, our study bridges the gap between healthy aging and MCI and dementia by including a still-healthy, but at-risk population, which is a population highlighted by the World Health Organization as a group most likely to benefit from preventive approaches aimed at delaying or preventing dementia [56]. Of note, the aim of our current study was to elucidate the mechanisms underlying maintaining optimal cognitive functioning in aging, as opposed to impacting mechanisms to prevent AD-incidence which requires longitudinal study designs incorporating different AD-related outcome measures.

Several limitations of the current study should be acknowledged. First, because participants were selected based on specific cardiovascular risk factors associated with a higher risk of future cognitive decline, it is possible that some individuals were already accumulating pathology that may lead to MCI or dementia in the future. Although the TICS-M1 was used during pre-screening, this instrument cannot completely guarantee to exclude early cognitive impairment. In future studies, blood biomarkers and cognitive assessment measures to exclude participants with a profile suggesting neurodegenerative diseases are warranted. This possibility complicates the interpretation of our findings with regard to the mechanisms involved in the preclinical phase of cognitive decline. Second, we used an active control group as opposed to a wait-list group. A wait-list design was considered unfeasible, as participants enrolling in a lifestyle intervention study already show motivation to improve aspects of their lifestyle, possibly even without the active guidance used in the intervention group. Assigning participants to a wait-list condition could be discouraging and might even result in higher drop-out rates. Additionally, due to ethical considerations, it was not possible to assign participants to a control group that required them to deliberately refrain from adhering to healthier lifestyle behaviours. As a result, we implemented two intervention arms that differed in intensity and level of guidance. The observed effect sizes of our neuroimaging and cognitive results might have been larger if we had included a control group not receiving generic lifestyle information. Because participants were included based on increased lifestyle-related risk factors associated with dementia incidence, resulting in generally unfavourable baseline risk profiles compared to the mean population, the observed improvements in both groups may be partially explained by regression to the mean. However, due to randomisation this attributed effect of regression to the mean would be expected to have a similar impact in both groups. Last, our study population had an unequal sex distribution (66% female, 34% male) and a relatively high education level (56% high educated), which may limit the generalizability of our findings to the broader population.

In conclusion, our findings indicate that older adults with modifiable lifestyle and cardiovascular risk factors that are associated with dementia risk improved their diet and sleep to a larger extent in the intervention group which received structured and supervised lifestyle intervention program, compared to the active control group which only received guidance by email on healthy living every two weeks. Other intervention domains, such as physical activity, stress management and wellbeing, and cognitive training did not show between-group differences. Within the intervention group, favourable reductions in lifestyle-modifiable cardiovascular risk factors, such as BMI and blood pressure, were observed, but these were not significantly different from changes in the active control group. No intervention effects were observed for brain and peripheral outcomes. Furthermore, we did find careful suggestions of a potential correlation between individual changes in CRP levels and CBF over time, emphasizing individual differences and the need for further research on the potential underlying mechanisms of neurobiological brain mechanisms.

## Data Availability

Data produced in the present study are available upon reasonable request to the authors.

## Acknowledgements

We wish to express our sincerest thanks to all participants for their valuable time and commitment to this study. We also wish to thank, remember and honour Wilma Steegenga, whose contributions as principal co-investigator have been invaluable in the design and practical implementation of the HELI study. We thank Noortje Overwater for her help in coordinating data collection at the Wageningen University & Research department of Human Nutrition and Health. We thank all involved members and staff of the HELI study, and all bachelor’s and master’s students who have helped with the practical aspects of conducting this study.

## Funding statement

This work was supported by a Crossover grant (MOCIA 17611) of the Dutch Research Council (NWO), granted in December 2019. The MOCIA program is a public-private partnership (see https://mocia.nl/scientific/).

## Conflicts of interest statement

JAHRC: PI in the ABOARD consortium. Local PI in EVOKE trial (NovoNordisk); consultancy or speaker for Eisai, Lilly, BMS, Novo Nordisk; Editor for JCBFM; SAB member for Alzheimer Nederland; Dutch National Guidelines committee member for dementia. IV: consultancy for Neurogen Biomarking, received research support for public-private partnerships (TKI, Health∼Holland) with Olink, Quanterix, Labonovum and Neurogen Biomarking, and performs contract research with Roche Diagnostics; all paid directly to her institution. All other authors declare no conflict of interest.

## Statement on informed written consent

All participants provided written informed consent to participate in the study.

## Notes

### Clinical Trial

NCT05777863

### Author Declarations

The Medical Research Ethics Committee (MREC) Oost-Nederland (file number NL78263.091.21) gave ethical approval for this work.

## References

1. Collaborators, G.D.F., Estimation of the global prevalence of dementia in 2019 and forecasted prevalence in 2050: an analysis for the Global Burden of Disease Study 2019. Lancet Public Health, 2022. 7(2): p. e105–e125.

2. Livingston, G., et al., Dementia prevention, intervention, and care: 2024 report of the Lancet standing Commission. Lancet, 2024. 404(10452): p. 572–628.

3. Livingston, G., et al., Dementia prevention, intervention, and care: 2020 report of the Lancet Commission. Lancet (London, England), 2020. 396(10248): p. 413–446.

4. Dhana, K., et al., Healthy lifestyle and the risk of Alzheimer dementia. Neurology, 2020. 95(4): p. e374.

5. Hankey, G.J., Public Health Interventions for Decreasing Dementia Risk. JAMA Neurology, 2018. 75(1): p. 11–12.

6. Norton, S., et al., Potential for primary prevention of Alzheimer’s disease: an analysis of population-based data. Lancet Neurol, 2014. 13(8): p. 788–94.

7. Ngandu, T., et al., A 2 year multidomain intervention of diet, exercise, cognitive training, and vascular risk monitoring versus control to prevent cognitive decline in at-risk elderly people (FINGER): a randomised controlled trial. Lancet, 2015. 385(9984): p. 2255–63.

8. Baker, L.D., et al., Structured vs Self-Guided Multidomain Lifestyle Interventions for Global Cognitive Function: The US POINTER Randomized Clinical Trial. Jama, 2025. 334(8): p. 681–691.

9. Andrieu, S., et al., Effect of long-term omega 3 polyunsaturated fatty acid supplementation with or without multidomain intervention on cognitive function in elderly adults with memory complaints (MAPT): a randomised, placebo-controlled trial. Lancet Neurol, 2017. 16(5): p. 377–389.

10. Demnitz-King, H., et al., Remote, lower-intensity, multidomain lifestyle intervention for subjective cognitive decline or mild cognitive impairment (APPLE-Tree): a multicentre, single-masked, randomised controlled trial. The Lancet Healthy Longevity, 2025. 6(10): p. 100777.

11. Heffernan, M., et al., Maintain Your Brain: Protocol of a 3-Year Randomized Controlled Trial of a Personalized Multi-Modal Digital Health Intervention to Prevent Cognitive Decline Among Community Dwelling 55 to 77 Year Olds. J Alzheimers Dis, 2019. 70(s1): p. S221–s237.

12. Beyer, F., et al., Exploring the effect of multi-modal intervention against cognitive decline on atrophy and small vessel disease imaging markers in the AgeWell.de imaging study. Neuroimage Clin, 2025. 46: p. 103796.

13. Crivelli, L., et al., Multidomain lifestyle intervention for the prevention of cognitive decline in at-risk older adults in Latin America (LatAm-FINGERS): a single-blind, multicentre, randomised controlled trial. The Lancet.

14. Meng, X., et al., Multidomain lifestyle interventions for cognition and the risk of dementia: A systematic review and meta-analysis. Int J Nurs Stud, 2022. 130: p. 104236.

15. Sakurai, T., et al., Japan-Multimodal Intervention Trial for the Prevention of Dementia: A randomized controlled trial. Alzheimer’s & Dementia, 2024. 20(6): p. 3918–3930.

16. Zülke, A.E., et al., A multidomain intervention against cognitive decline in an at-risk-population in Germany: Results from the cluster-randomized AgeWell.de trial. Alzheimer’s & Dementia, 2024. 20(1): p. 615–628.

17. van Loenen, M.R., et al., The Effects of a Multidomain Lifestyle Intervention on Brain Function and Its Relation With Immunometabolic Markers and Intestinal Health in Older Adults at Risk of Cognitive Decline: Study Design and Baseline Characteristics of the HELI Randomized Controlled Trial. JMIR Res Protoc, 2025. 14.

18. Sindi, S., et al., The CAIDE Dementia Risk Score App: The development of an evidence-based mobile application to predict the risk of dementia. Alzheimers Dement (Amst), 2015. 1(3): p. 328–33.

19. Bull, F.C., et al., World Health Organization 2020 guidelines on physical activity and sedentary behaviour. Br J Sports Med, 2020. 54(24): p. 1451–1462.

20. van den Berg, E., et al., The Telephone Interview for Cognitive Status (Modified): relation with a comprehensive neuropsychological assessment. J Clin Exp Neuropsychol, 2012. 34(6): p. 598–605.

21. Seblova, D., R. Berggren, and M. Lövdén, Education and age-related decline in cognitive performance: Systematic review and meta-analysis of longitudinal cohort studies. Ageing Res Rev, 2020. 58: p. 101005.

22. Deckers, K., et al., A multidomain lifestyle intervention to maintain optimal cognitive functioning in Dutch older adults-study design and baseline characteristics of the FINGER-NL randomized controlled trial. Alzheimers Res Ther, 2024. 16(1): p. 126.

23. Kivipelto, M., et al., World-Wide FINGERS Network: A global approach to risk reduction and prevention of dementia. Alzheimers Dement, 2020. 16(7): p. 1078–1094.

24. van Loenen, M.R., et al., Integrating lifestyle, vascular, brain and cognitive markers: an exploratory multimodal approach to brain aging. medRxiv, 2026: p. 2026.01.08.26343661.

25. van Lee, L., et al., Evaluation of a screener to assess diet quality in the Netherlands. Br J Nutr, 2016. 115(3): p. 517–26.

26. Morris, M.C., et al., MIND diet slows cognitive decline with aging. Alzheimers Dement, 2015. 11(9): p. 1015–22.

27. Beers, S., et al., Development of the Dutch Mediterranean-Dietary Approaches to Stop Hypertension Intervention for Neurodegenerative Delay (MIND) Diet and its scoring system, alongside the modification of a brief FFQ for assessing dietary adherence. Br J Nutr, 2026. 135(5): p. 528–536.

28. Wendel-Vos, G.C.W., et al., Reproducibility and relative validity of the short questionnaire to assess health-enhancing physical activity. Journal of Clinical Epidemiology, 2003. 56(12): p. 1163–1169.

29. Rosenberg, D.E., et al., Reliability and validity of the Sedentary Behavior Questionnaire (SBQ) for adults. J Phys Act Health, 2010. 7(6): p. 697–705.

30. Lee, E.H., Review of the psychometric evidence of the perceived stress scale. Asian Nurs Res (Korean Soc Nurs Sci), 2012. 6(4): p. 121–7.

31. Zigmond, A.S. and R.P. Snaith, The hospital anxiety and depression scale. Acta Psychiatr Scand, 1983. 67(6): p. 361-70.

32. Baer, R.A., et al., Construct validity of the five facet mindfulness questionnaire in meditating and nonmeditating samples. Assessment, 2008. 15(3): p. 329–342.

33. Buysse, D.J., et al., The Pittsburgh Sleep Quality Index: a new instrument for psychiatric practice and research. Psychiatry Res, 1989. 28(2): p. 193–213.

34. Nath Kundu, R., S. Biswas, and M. Das, Mean Arterial Pressure Classification: A Better Tool for Statistical Interpretation of Blood Pressure Related Risk Covariates. Cardiology and Angiology: An International Journal, 2017. 6(1): p. 1–7.

35. Bowie, C.R. and P.D. Harvey, Administration and interpretation of the Trail Making Test. Nat Protoc, 2006. 1(5): p. 2277–81.

36. Carone, D.A., E. Strauss, E. M. S. Sherman, & O. Spreen, A Compendium of Neuropsychological Tests: Administration, Norms, and Commentary. Applied Neuropsychology, 2007. 14(1): p. 62–63.

37. Ryan, J.J. and S.J. Lopez. Wechsler Adult Intelligence Scale-III. 2001.

38. Rey, A., L’examen clinique en psychologie. 1964: Presses universitaires de France.

39. Verberk, I.M.W., et al., Development of thresholds and a visualization tool for use of a blood test in routine clinical dementia practice. Alzheimers Dement, 2024. 20(9): p. 6115–6132.

40. Kruger, K., et al., Evaluation of inter- and intra-variability in gut health markers in healthy adults using an optimised faecal sampling and processing method. Scientific Reports, 2024. 14(1): p. 24580.

41. Bolyen, E., et al., Reproducible, interactive, scalable and extensible microbiome data science using QIIME 2. Nat Biotechnol, 2019. 37(8): p. 852–857.

42. McMurdie, P.J. and S. Holmes, Phyloseq: a bioconductor package for handling and analysis of high-throughput phylogenetic sequence data. Pac Symp Biocomput, 2012: p. 235–46.

43. Kembel, S.W., et al., Picante: R tools for integrating phylogenies and ecology. Bioinformatics, 2010. 26(11): p. 1463–1464.

44. Benjamini, Y. and Y. Hochberg, Controlling the false discovery rate: a practical and powerful approach to multiple testing. Journal of the Royal statistical society: series B (Methodological), 1995. 57(1): p. 289–300.

45. Ishii, S., et al., Gender, obesity and repeated elevation of C-reactive protein: data from the CARDIA cohort. PLoS One, 2012. 7(4): p. e36062.

46. Remie, L.B., et al., MIND-NL diet adherence moderates the relation of low-grade systemic inflammation with neuroinflammatory metabolites and cognitive functioning: an exploratory cross-sectional study in older adults. J Neuroinflammation, 2026. 23(1).

47. Baart, A.M., et al., Assessment of the Dutch Healthy Diet index 2015 in the Lifelines cohort study at baseline. Eur J Clin Nutr, 2024. 78(3): p. 217–227.

48. Li, W., et al., Differential developmental trajectories of magnetic susceptibility in human brain gray and white matter over the lifespan. Hum Brain Mapp, 2014. 35(6): p. 2698–713.

49. Nijssen, K.M.R., et al., Longer-term mixed nut consumption improves brain vascular function and memory: A randomized, controlled crossover trial in older adults. Clin Nutr, 2023. 42(7): p. 1067–1075.

50. Kleinloog, J.P.D., et al., Aerobic Exercise Training Improves Cerebral Blood Flow and Executive Function: A Randomized, Controlled Cross-Over Trial in Sedentary Older Men. Front Aging Neurosci, 2019. 11: p. 333.

51. Thomas, B.P., et al., Brain Perfusion Change in Patients with Mild Cognitive Impairment After 12 Months of Aerobic Exercise Training. J Alzheimers Dis, 2020. 75(2): p. 617–631.

52. Stephen, R., et al., Brain volumes and cortical thickness on MRI in the Finnish Geriatric Intervention Study to Prevent Cognitive Impairment and Disability (FINGER). Alzheimers Res Ther, 2019. 11(1): p. 53.

53. Bowtell, J.L., et al., Enhanced task-related brain activation and resting perfusion in healthy older adults after chronic blueberry supplementation. 2017. 42(7): p. 773–779.

54. Colcombe, S.J., et al., Cardiovascular fitness, cortical plasticity, and aging. Proc Natl Acad Sci U S A, 2004. 101(9): p. 3316–21.

55. Petersen, R.C., et al., Current concepts in mild cognitive impairment. Arch Neurol, 2001. 58(12): p. 1985–92.

56. World Health Organization, Risk reduction of cognitive decline and dementia: WHO guidelines. 2019: Geneva. p. 96.

