## Supplementary Files for "The Effects of a Multidomain Lifestyle Intervention on Brain Function and Its Relation with Immunometabolic Markers and Intestinal Health in Older Adults at Risk of Cognitive Decline: The HELI Randomised Controlled Trial"

**Supplementary File 1.** Preprocessing and analysis of neuroimaging measures.

Image preprocessing of the T1-weighted MP2RAGE images included cortical reconstruction and volumetric segmentation using FreeSurfer's aparc+aseg pipeline (http://surfer.nmr.mgh.harvard.edu/). From the resulting parcellations, whole-brain grey matter (GM), white matter (WM), and hippocampal masks were derived from the ASEG atlas (aseg_stats), while the rostral middle frontal region from the Desikan-Kiliany atlas (aparc_volume) was used as a proxy for the dlPFC. Volumes of whole-brain GM, WM, dlPFC and hippocampus were subsequently derived from the ASEG parcellations. All volumetric measures were calculated to mm3.
Preprocessing of the fMRI images was performed with fMRIPrep (23.2.0; RRID:SCR_016216), including realignment, distortion correction, coregistration to native anatomical scans, and normalization to MNI152 space. Prior to analysis, preprocessed fMRI images were spatially smoothed with a 6-mm full-width at half-maximum (FWHM) Gaussian kernel using Statistical Parametric Mapping 12 (SPM12; https://www.fil.ion.ucl.ac.uk/spm/software/spm12/), implemented in MATLAB R2024a (Mathworks inc.; https://nl.mathworks.com/products/matlab.html). Using generalized linear models in SPM12, first-level analyses were performed with regressors modelling the three task conditions. Confound regressors per session included motion parameters, framewise displacement, six aCompCor and tCompCor components, every cosine regressor, and ICA-AROMA noise components (excluding those correlating >0.05 with the task design). The main working memory contrast of interest was 2-back minus 0-back. For the ROI analyses, the dlPFC mask was created by combining working memory-related brain activation from a Neurosynth [1, 2] map with peak dlPFC voxels in a prior meta-analysis on aging-related N-back effects [3]. The hippocampus mask was derived from the Harvard-Oxford subcortical atlas [4]. We used the mean β-values within our ROIs for the 2-back minus 0-back (2b-0b) contrast, derived from SPM12, as part of the group analyses.

The Hadamard time-encoded pCASL perfusion images were preprocessed using the FSL Linear Image Registration Tool (FLIRT) and subsequently decoded using a Hadamard decoding scheme to acquire motion-corrected decoded perfusion images. We used the FSL BASIL 'asl_gui' toolbox to generate quantified whole-brain CBF maps. We used BASIL to analyze 14 pre-subtracted perfusion volumes (7 post-labelling delays with 2 repeats) using a 0.4 second bolus duration (0.5 – 2.9 s) and a 3D GRASE readout. For anatomical reference and registration of the CBF maps, the T1-weighted MP2RAGE scan was processed with fsl_anat, and the corresponding output was supplied to BASIL. Calibration was performed with a proton-density M0 calibration image (TR/TE = 3000/16.28 ms, flip angle = 120°, voxel size = 1.8×1.8×3.5 mm), and an ASL-specific mask was provided to only assess voxels containing perfusion signals. GM, WM, and hippocampus masks were derived from FreeSurfer ASEG parcellations, while the standard space dlPFC mask, initially created for the fMRI analysis, was transformed to native space. Quantified CBF values were subsequently extracted from the voxels of each respective mask. Averaged CBF values were then calculated across all voxel CBF values within each mask.

We used intracranial myo-inositol concentrations (with total water as a reference), measured with MRS, as a marker of neuroinflammation [5]. Myo-inositol concentrations were quantified within a 20mm isotropic voxel positioned in the left dlPFC. Spectra were acquired with a PRESS sequence in combination with chemically selective water suppression (CHESS). Voxel placement was manually guided by the participant's T1-weighted image, whilst avoiding non-brain tissue such as CSF, meninges or the skull. Before acquisition, shimming was performed within the voxel to minimize local field inhomogeneity. Preprocessing of both water-suppressed and unsuppressed spectra was conducted in Osprey 2.4.0 software. By preprocessing metabolite and lineshape reference, as well as native anatomical T1-weighted MP2RAGE images, averaged spectra were created. If necessary, water-unsuppressed lineshape reference data was used to perform eddy-current correction [6]. Metabolite spectra were modeled over 0.5–4.0 ppm (with 0.4 ppm knot spacing). Water reference spectra were fitted separately over 2.0–7.4 ppm. After coregistration of the spectroscopy data to the T1-weighted image, we performed tissue segmentation in SPM12 to estimate total GM, WM and CSF fractions within the voxel. Total metabolite estimates were corrected for tissue composition, relaxation times, and scaled to total water concentration. Final myo-inositol estimates were additionally adjusted for the spectral quality measure of relative residual amplitude (relResA).

**References:**


1. Yarkoni, T., et al., *Large-scale automated synthesis of human functional neuroimaging data.* Nature Methods, 2011. **8**(8): p. 665-670.
2. Yarkoni, T., et al. *Working Memory: An automated meta-analysis of 1091 studies*. [cited 2025; Available from: https://neurosynth.org/analyses/terms/working%20memory/.
3. Wang, H., et al., *A coordinate-based meta-analysis of the n-back working memory paradigm using activation likelihood estimation.* Brain Cogn, 2019. **132**: p. 1-12.
4. Jenkinson, M., et al., *FSL.* Neuroimage, 2012. **62**(2): p. 782-90.
5. Chang, L., et al., *Magnetic resonance spectroscopy to assess neuroinflammation and neuropathic pain.* J Neuroimmune Pharmacol, 2013. **8**(3): p. 576-93.
6. Klose, U., *In vivo proton spectroscopy in presence of eddy currents.* Magn Reson Med, 1990. **14**(1): p. 26-30.

**Supplementary File 2**. The within-group difference of the lifestyle intervention domains.

| | Intervention group | | | | | | | Active control group | | | | | | |
|---|---|---|---|---|---|---|---|---|---|---|---|---|---|---|
| | Baseline | | Six-month | | Within-group difference | | | Baseline | | Six-month | | Within-group difference | | |
| | n | Mean ±SD | n | Mean ±SD | Mean difference, SE | Unadjusted P-values | FDR adjusted P-values[1] | n | Mean ±SD | n | Mean ±SD | Mean difference, SE | Unadjusted P-values | FDR adjusted P-values[1] |
| Lifestyle questionnaires | | | | | | | | | | | | | | |
| DHD-index (/150) | 53 | 100 ±18 | 47 | 112 ±19 | 11.42, 2.24 | <0.0001 | **<0.0001** | 49 | 101 ±16 | 39 | 106 ±17 | 4.73, 2.44 | 0.06 | 0.24 |
| MIND-NL (/15) | 53 | 8.4 ±2.2 | 46 | 10.5 ±2.2 | 2.07, 0.36 | <0.0001 | **<0.0001** | 47 | 8.7 ±1.9 | 36 | 9.1 ±1.7 | 0.40, 0.40 | 0.33 | 0.44 |
| SQUASH (min/week) | 53 | 6300 ±3710 | 47 | 7400 ±4660 | 1050, 552 | 0.06 | 0.10 | 48 | 6290 ±3390 | 36 | 6990 ±4810 | 752, 626 | 0.23 | 0.38 |
| LASA-SBQ (min/day) | 53 | 462 ±219 | 47 | 502 ±201 | 39.2, 35.0 | 0.27 | 0.36 | 49 | 493 ±205 | 37 | 479 ±249 | -11.8, 38.9 | 0.76 | 0.76 |
| PSS (/40) | 53 | 15.1 ±5.0 | 47 | 14.9 ±5.6 | 0.09, 0.57 | 0.87 | 0.87 | 49 | 15.1 ±4.8 | 38 | 13.9 ±5.4 | -1.04, 0.63 | 0.10 | 0.27 |
| HADS (/42) | 53 | 7.5 ±5.4 | 47 | 6.2 ±4.7 | -1.30, 0.60 | 0.03 | 0.06 | 49 | 7.3 ±4.8 | 37 | 7.0 ±4.9 | -0.30, 0.67 | 0.66 | 0.75 |
| FFMQ (/195) | 53 | 87.3 ±7.8 | 48 | 86.7 ±8.5 | -0.47, 0.95 | 0.63 | 0.72 | 49 | 87.4 ±10.0 | 38 | 89.4 ±9.7 | 2.03, 1.06 | 0.06 | 0.24 |
| PSQI (/21) | 53 | 6.7 ±3.6 | 47 | 5.3 ±3.0 | -1.36, 0.34 | 0.0001 | **0.0003** | 49 | 5.4 ±3.1 | 37 | 5.6 ±3.3 | 0.45, 0.38 | 0.24 | 0.38 |

| | | | | | | | | | | | | |
|---|---|---|---|---|---|---|---|---|---|---|---|---|
| Cognitive assessment | | | | | | | | | | | | |
| TMT (TMT-B/TMT-A score) | 53 | 2.31 ±0.82 | 47 | 2.20 ±0.63 | -0.11, 0.10 | 0.28 | 0.47 | 49 | 2.17 ±0.55 | 39 | 2.20 ±0.70 | 0.02, 0.11 | 0.87 | 0.87 |
| VFT (total score) | 53 | 26.0 ±6.06 | 47 | 26.1 ±5.31 | 0.33, 0.64 | 0.61 | 0.61 | 49 | 26.0 ±4.89 | 39 | 26.6 ±5.69 | 0.53, 0.70 | 0.45 | 0.56 |
| DSST (total score) (/90) | 53 | 52.6 ±9.70 | 47 | 54.9 ±10.0 | 2.14, 0.82 | 0.01 | **0.03** | 49 | 51.3 ±10.6 | 39 | 53.1 ±10.3 | 2.25, 0.90 | 0.01 | **0.03** |
| RAVLT delayed recall (/15) | 53 | 7.77 ±3.24 | 47 | 10.3 ±3.21 | 2.32, 0.39 | <0.0001 | **<0.0001** | 49 | 7.51 ±3.12 | 39 | 9.59 ±3.36 | 2.14, 0.43 | <0.0001 | **<0.0001** |
| WAIS digit span (/28) | 53 | 15.5 ±3.69 | 47 | 15.4 ±3.77 | -0.29, 0.34 | 0.41 | 0.51 | 49 | 14.9 ±3.10 | 39 | 15.3 ±3.47 | 0.67, 0.37 | 0.08 | 0.13 |

[1] FDR correction per group (intervention and active control) and per subgroup outcomes.

Abbreviations: FFMQ, Five Facet Mindfulness Questionnaire; LASA, Sedentary Behaviour Questionnaire; DHD, Dutch healthy diet; PSS, Perceived Stress Scale; PSQI, Pittsburgh Sleep Quality Index; SQUASH, Short Questionnaire to Assess Health-enhancing physical activity; HADS, Hospital and Anxiety Depression Scale.

**Supplementary File 3.** The within-group differences of the primary outcomes.

| | Intervention group | | | | | | | Active control group | | | | | | |
|---|---|---|---|---|---|---|---|---|---|---|---|---|---|---|
| | Baseline [1] | | Six-month | | Within-group difference | | | Baseline | | Six-month | | Within-group difference | | |
| | n | Mean ±SD | n | Mean ±SD | Mean difference, SE | Unadjusted P-values | FDR adjusted P-values [1] | n | Mean ±SD | n | Mean ±SD | Mean difference, SE | Unadjusted P-values | FDR adjusted P-values [1] |
| Working memory fMRI BOLD response in dlPFC (2b-0b) | 50 | 0.45 ±0.13 | 40 | 0.38 ±0.15 | 0.04, 0.02 | 0.08 | 0.17 | 45 | 0.37 ±0.15 | 37 | 0.41 ±0.18 | 0.04, 0.03 | 0.15 | 0.33 |
| Working memory fMRI BOLD response in hippocampus (2b-0b) | 50 | -0.02 ±0.06 | 40 | -0.05 ±0.07 | -0.03, 0.01 | 0.07 | 0.17 | 45 | -0.04 ±0.08 | 37 | -0.05 ±0.10 | -0.006, 0.01 | 0.69 | 0.76 |
| Working memory N-back task accuracy (d' 2b) | 51 | 2.31 ±0.87 | 43 | 2.53 ±0.88 | 0.23, 0.11 | 0.04 | 0.15 | 46 | 2.36 ±0.62 | 38 | 2.55 ±0.65 | 0.23, 0.12 | 0.048 | 0.18 |
| CBF in dlPFC [2] (ml/100g tissue/min) | 50 | 40.0 ±13.6 | 40 | 42.2 ±12.1 | 1.79, 1.66 | 0.28 | 0.31 | 44 | 37.6 ±11.4 | 34 | 43.6 ±11.6 | 6.39, 1.84 | 0.0008 | **0.009** |
| CBF in hippocampus [2] (ml/100g tissue/min) | 50 | 42.2 ±12.1 | 40 | 40.7 ±9.14 | -2.36, 1.57 | 0.14 | 0.22 | 44 | 40.8 ±10.6 | 34 | 44.8 ±7.95 | 4.26, 1.74 | 0.02 | 0.11 |
| Plasma hs-CRP levels (mg/L) | 47 | 2.97 ±3.10 | 47 | 3.76 ±4.88 | 0.79, 0.46 | 0.09 | 0.17 | 38 | 2.78 ±2.87 | 38 | 1.89 ±1.38 | -0.91, 0.51 | 0.08 | 0.22 |
| Plasma IL-6 levels (pg/ml) | 45 | 0.88 ±0.60 | 45 | 0.99 ±0.91 | 0.11, 0.11 | 0.31 | 0.31 | 37 | 0.94 ±0.66 | 37 | 0.81 ±0.57 | -0.13, 0.12 | 0.30 | 0.47 |

| | | | | | | | | | | | | | |
|---|---|---|---|---|---|---|---|---|---|---|---|---|---|
| Plasma TNF-α levels (pg/ml) | 45 | 1.22 ±0.49 | 45 | 1.39 ±1.32 | 0.17, 0.14 | 0.22 | 0.30 | 37 | 1.25 ±0.32 | 37 | 1.22 ±0.42 | -0.03, 0.15 | 0.86 | 0.86 |
| Microbiota Shannon diversity | 47 | 4.1 ± 0.5 | 47 | 4.2 ± 0.5 | 0.08, 0.07 | 0.29 | 0.31 | 39 | 4.1 ± 0.7 | 39 | 4.2 ± 0.6 | 0.07, 0.07 | 0.34 | 0.47 |
| Microbiota phylogenetic diversity | 47 | 21.8 ± 5.7 | 47 | 22.7 ± 6.0 | 0.90, 0.37 | 0.02 | 0.11 | 39 | 21.2 ± 5.2 | 39 | 21.7 ± 5.2 | 0.40, 0.40 | 0.32 | 0.47 |
| Microbiota Chao1 richness | 47 | 237.9 ± 86.5 | 47 | 252.6 ± 93.1 | 14.70, 4.86 | 0.003 | **0.03** | 39 | 228.3 ± 85.3 | 39 | 232.0 ± 82.1 | 3.73, 5.33 | 0.49 | 0.60 |

[1] FDR correction per group (intervention and active control)

[2] model adjusted for pCASL sequence version (pre- vs. post-MRI scanner update)

Abbreviations: dlPFC, dorsolateral prefrontal cortex; CBF, cerebral blood flow.

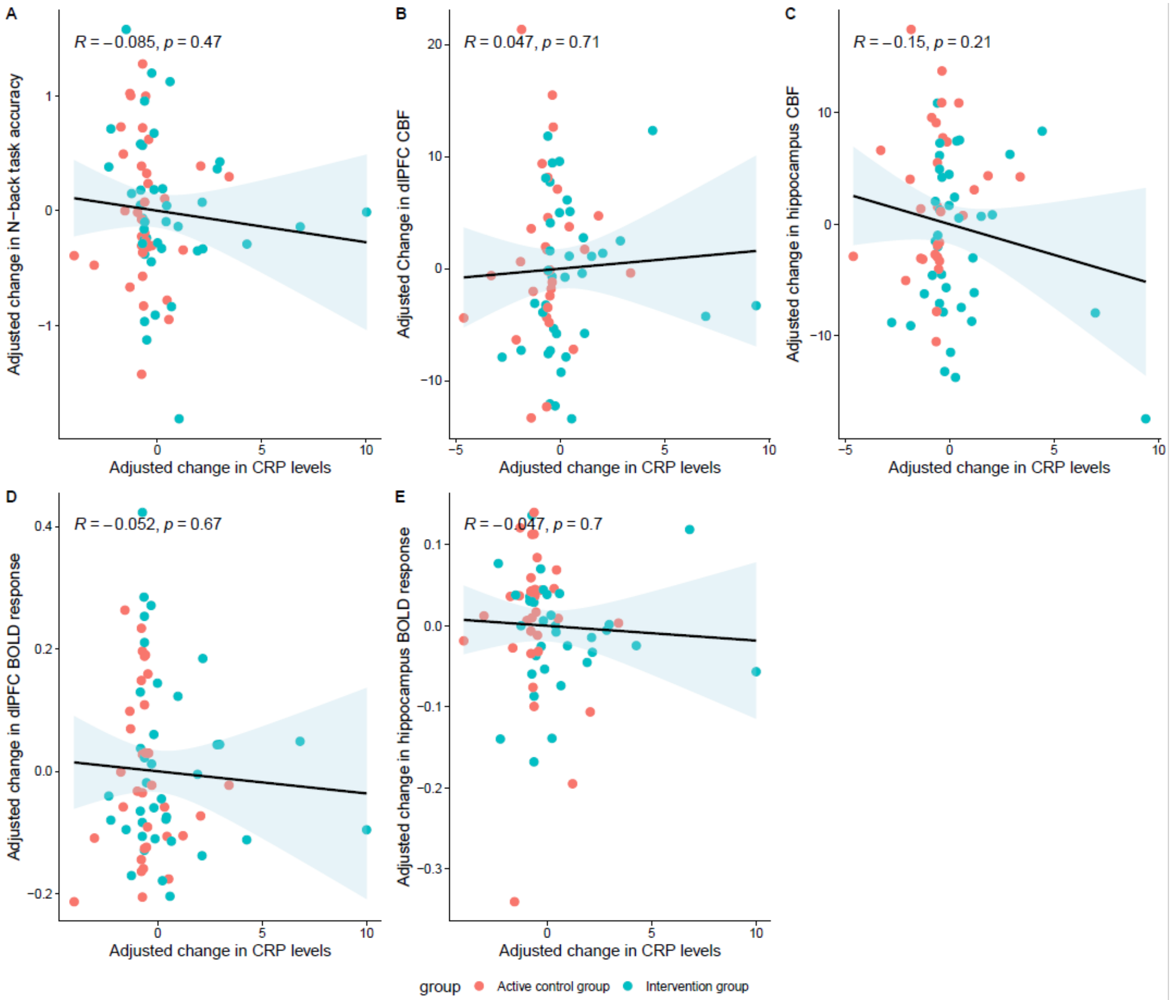


**Supplementary File 4.** The correlation between changes in CRP levels and (A) change in N-back task accuracy, (B) change in CBF in dlPFC, (C) change in CBF in hippocampus, (D) change in fMRI BOLD responses, (E) change in fMRI BOLD responses **after excluding participants with acute infections**. Changes were adjusted for their baseline levels before calculation of the Pearson correlation. Also, the CBF measures were adjusted for the pCASL sequence. Data points are colored by intervention group. The solid line indicates the fitted regression line and the shaded area represents the 95% confidence interval. P-values are not FDR corrected.

**Supplementary File 5.** The within-group differences of the (other) secondary outcomes.

| | Intervention group | | | | | | | Active control group | | | | | | |
|---|---|---|---|---|---|---|---|---|---|---|---|---|---|---|
| | Baseline | | Six-month | | Within-group difference | | | Baseline | | Six-month | | Within-group difference | | |
| | n | Mean ±SD | n | Mean ±SD | Mean difference, SE | Unadjusted P-values | FDR adjusted P-values [1] | n | Mean ±SD | n | Mean ±SD | Mean difference, SE | Unadjusted P-values | FDR adjusted P-values [1] |
| Anthropometric measurements | | | | | | | | | | | | | | |
| BMI (kg/m$^2$) | 53 | 31.0 ±5.9 | 47 | 30.3 ± 5.7 | -0.51, 0.21 | 0.02 | **0.02** | 49 | 28.9 ±4.7 | 39 | 28.4 ±4.7 | 0.001, 0.23 | 1.00 | 1.00 |
| Waist-hip ratio | 53 | 0.97 ±0.1 | 47 | 0.93 ±0.1 | -0.04, 0.01 | 0.005 | **0.008** | 49 | 0.94 ±0.1 | 39 | 0.93 ±0.1 | -0.008, 0.01 | 0.59 | 0.89 |
| Blood pressure (mmHg) | 52 | 110 ±14 | 46 | 105 ±11 | -5.19, 1.32 | 0.0002 | **0.0006** | 49 | 107 ±13 | 39 | 104 ±12 | -2.10, 1.43 | 0.15 | 0.45 |
| Cardio-metabolic profile | | | | | | | | | | | | | | |
| Glucose (mmol/L) | 53 | 6.7 ±2.5 | 46 | 6.3 ±1.7 | -0.35, 1.19 | 0.07 | 0.11 | 48 | 6.3 ±1.3 | 36 | 6.2 ±1.6 | 0.03, 0.22 | 0.90 | 1.00 |
| Insulin (mmol/L) | 49 | 24.7 ±58.2 | 46 | 24.3 ±60.1 | -0.09, 0.76 | 0.91 | 0.91 | 45 | 13.6 ±8.8 | 36 | 12.9 ±8.5 | -0.63, 0.86 | 0.46 | 1.00 |
| Cholesterol (mmol/L) | 53 | 5.5 ±1.0 | 46 | 5.2 ±1.0 | -0.33, 0.11 | 0.004 | **0.01** | 48 | 5.7 ±1.3 | 37 | 5.7 ±1.3 | -0.08, 0.13 | 0.53 | 1.00 |
| HDL-cholesterol (mmol/L) | 53 | 1.5 ±0.4 | 46 | 1.5 ±0.4 | -0.01, 0.03 | 0.68 | 0.82 | 48 | 1.4 ±0.4 | 37 | 1.5 ±0.4 | 0.3e-4 | 1.00 | 1.00 |

| | | | | | | | | | | | | | | |
|---|---|---|---|---|---|---|---|---|---|---|---|---|---|---|
| LDL-cholesterol (mmol/L) | 53 | 3.3 ±0.9 | 46 | 3.0 ±0.8 | -0.28, 0.10 | 0.005 | **0.01** | 48 | 3.5 ±1.1 | 37 | 3.5 ±1.0 | -0.04, 0.11 | 0.69 | 1.00 |
| Triglycerides (mmol/L) | 53 | 1.7 ±0.7 | 46 | 1.6 ±0.7 | -0.18, 0.06 | 0.006 | **0.01** | 48 | 1.6 ±0.8 | 37 | 1.6 ±0.7 | 0.04, 0.07 | 0.54 | 1.00 |
| Neuroimaging | | | | | | | | | | | | | | |
| Hippocampal volume [2] ($mm^3$) | 53 | 7870 ±765 | 45 | 7830 ±702 | -44.1, 24.9 | 0.08 | 0.24 | 48 | 7760 ±777 | 38 | 7720 ±763 | -37.8, 27.1 | 0.17 | 0.51 |
| dlPFC volume [2] ($mm^3$) | 53 | 24500 ±2830 | 45 | 24600 ±3030 | 9.84, 153 | 0.95 | 0.95 | 48 | 23700 ±2930 | 38 | 23600 ±2580 | -27.7, 166 | 0.87 | 0.87 |
| Brain myo-inositol levels (mol/kg) | 52 | 10.3 ±3.4 | 45 | 9.8 ±1.3 | -0.50, 0.47 | 0.29 | 0.44 | 48 | 10.4 ±1.9 | 36 | 10.6 ±2.5 | 0.16, 0.51 | 0.76 | 0.87 |
| Inflammation profile | | | | | | | | | | | | | | |
| IFN-γ (pg/mL) | 45 | 4.49 ±2.48 | 45 | 9.44 ±28.9 | 4.96, 3.34 | 0.14 | 0.26 | 37 | 6.17 ±7.59 | 37 | 5.51 ±5.31 | 0.65, 3.68 | 0.86 | 0.93 |
| IL-8 (pg/mL) | 45 | 4.57 ±1.30 | 45 | 4.54 ±1.56 | -0.03, 0.20 | 0.90 | 0.90 | 37 | 4.70 ±1.18 | 37 | 4.75 ±1.69 | 0.04, 0.22 | 0.84 | 0.93 |
| IL-10 (pg/mL) | 45 | 0.31 ±0.79 | 45 | 0.69 ±3.27 | 0.38, 0.28 | 0.17 | 0.26 | 37 | 0.28 ±0.19 | 37 | 0.25 ±0.15 | 0.03, 0.30 | 0.93 | 0.93 |
| Intestinal integrity | | | | | | | | | | | | | | |
| LBP (μg/ml) | 39 | 13.7 ±4.1 | 38 | 13.9 ±4.4 | 0.53, 0.44 | 0.24 | 0.36 | 36 | 12.8 ±3.7 | 35 | 12.8 ±3.5 | -0.59, 0.50 | 0.24 | 0.36 |

| | | | | | | | | | | | | | |
|---|---|---|---|---|---|---|---|---|---|---|---|---|---|
| LPS ( EU/ml) | 39 | 4.3 ±13.7 | 38 | 26.2 ±109.0 | 16.59, 13.8 | 0.23 | 0.36 | 36 | 17.8 ±65.9 | 35 | 19.1 ±59.7 | 0.52, 15.5 | 0.97 | 0.97 |
| Zonulin (ng/ml) | 39 | 40.3 ±16.7 | 38 | 39.3 ±17.0 | 0.05, 1.20 | 0.97 | 0.97 | 36 | 37.2 ±17.0 | 35 | 36.0 ±15.1 | -1.97, 1.36 | 0.15 | 0.36 |
| Biomarkers of neurodegeneration | | | | | | | | | | | | | | |
| NfL (pg/mL) | 47 | 15.0 ±6.1 | 46 | 15.0 ±5.6 | 0.01, 0.37 | 0.97 | 0.97 | 38 | 13.1 ±5.5 | 36 | 14.2 ±6.2 | 1.10, 0.42 | 0.01 | **0.02** |
| Aβ42/40 ratio | 47 | 0.057 ±0.011 | 46 | 0.057 ±0.012 | -0.0002, 0.0007 | 0.73 | 0.97 | 38 | 0.058 ±0.011 | 36 | 0.058 ±0.011 | 0.0009, 0.0008 | 0.25 | 0.25 |
| GFAP (pg/mL) | 47 | 77.0 ±24.7 | 46 | 80.0 ±29.5 | 3.26, 1.74 | 0.07 | 0.21 | 38 | 71.9 ±21.4 | 36 | 79.1 ±24.7 | 7.03, 197 | 0.0006 | **0.002** |
| Other | | | | | | | | | | | | | | |
| White blood cell count (counts×$10^9$/L) | 53 | 6.2 ±1.9 | 47 | 6.5 ±2.5 | 0.36, 0.28 | 0.20 | - | 49 | 6.3 ±1.8 | 38 | 5.2 ±1.8 | -0.09, 0.31 | 0.76 | - |

[1] FDR correction per group (intervention and active control) and per subgroup outcomes.

[2] Model adjusted for total brain volume.
